# Pro-cognitive effects of 5-HT4 receptor agonism in individuals with remitted depression

**DOI:** 10.1101/2025.08.20.25332371

**Authors:** A.N. de Cates, S. Hamilton, A. Guru, M. Blandhol, M. Colwell, P.J. Cowen, M. Simmons, B. Jones, C.J. Harmer, S.E. Murphy

## Abstract

**Background:** Cognitive impairment is a common and persistent feature of depression, yet remains poorly understood and inadequately treated. Preclinical and human studies suggest that stimulating 5-HT_4_ receptors (5-HT_4_R) enhances neuroplasticity and rapidly improves learning and memory. This study is the first to examine the cognitive effects of 5-HT_4_R agonism in adults with a history of recurrent depression. We hypothesised that short term 5-HT_4_R agonist administration would produce a broad profile of pro-cognitive effects.

**Methods:** 50 participants not currently depressed but with at least two previous episodes of depression (remitted depression) were randomised in a double-blind design to receive either prucalopride (2mg daily titrated from 1mg over 2 days) or placebo for 7-10 days. Participants completed self-report questionnaires and a cognitive task battery assessing declarative memory, working memory, emotional processing, and executive function before and after medication.

**Results:** Compared to placebo, prucalopride significantly improved word recall on an auditory verbal learning task, and was associated with faster response times on a complex working memory task without loss of accuracy. It also improved the accurate recognition of rapidly presented facial expressions. Prucalopride had minimal effects on emotionally-valenced cognitive tasks, consistent with previous findings. Cognitive improvements were independent of baseline mood symptoms or self-reported cognitive difficulties.

**Conclusions:** Short-term 5-HT_4_R agonism improved performance on multiple objective cognitive measures in individuals with a history of depression. These findings replicate our previous results in healthy volunteers using prucalopride and support a role for 5-HT_4_Rs as a promising target for cognitive enhancement in mood disorders.

## Introduction

Cognitive impairment is a common feature of depression, with significant personal and societal costs. These symptoms are poorly addressed by first-line antidepressant treatments (Millan et al., 2012). Around 80% of individuals experience cognitive difficulties during a current episode of depression (Conradi, Ormel, & de Jonge, 2011), and over 40% continue to report subjective cognitive problems even after other symptoms of depression have resolved (McClintock et al., 2011; Rock, Roiser, Riedel, & Blackwell, 2014; Semkovska et al., 2019). Despite their prevalence and impact on functional recovery, treatment options targeting cognitive dysfunction in depression remain limited (Halahakoon & Roiser, 2016). Moreover, persistent cognitive deficits following remission may serve as a risk marker for future relapse (Halahakoon, Lewis, & Roiser, 2019), highlighting the need for novel therapeutic strategies to address cognitive symptoms and support long-term recovery.

One promising target is the serotonin 4 receptor (5-HT_4_R). A robust body of preclinical evidence demonstrates that 5-HT_4_R agonists rapidly improve performance on tasks of learning and memory (Fontana, Daniels, Wong, Clark, & Eglen, 1997; Hagena & Manahan-Vaughan, 2017; King, Marsden, ^&^ Fone, 2008^;^ Lamirault & Simon, 2001; Marchetti et al., 2011). This effects are blocked by 5-HT_4_R antagonists, indicating a direct receptor-mediated mechanism (Fontana et al., 1997). There are a number of potential mechanisms of these pro-cognitive effects, including enhanced hippocampal synaptic plasticity (Kozono, Ohtani, & Shiga, 2017), increased acetylcholine release (Siniscalchi, Badini, Beani, & Bianchi, 1999), and modulation of glutamate transmission (Chen et al., 2020).

Early translational studies in healthy volunteers (de Cates et al., 2022; de Cates et al., 2021; S. ^M^urphy, Wright, Browning, Cowen, & Harmer, 2020^b^) have demonstrated that the selective 5-HT_4_R agonist prucalopride has a pro-cognitive profile in humans. In these studies, acute and subchronic (6 days) administration of prucalopride has been shown to improve learning and memory across a range of cognitive tasks, including declarative memory and reward learning. Prucalopride also improves the accuracy of task performance in paradigms without a memory component, which may be indicative of enhanced attentional function. For example, prucalopride has been shown to increase the accuracy with which healthy volunteers are able to identify the gender of rapidly presented faces (de Cates et al., 2022). Consistent with these pro-cognitive behavioural effects, neuroimaging data show prucalopride increases bilateral hippocampal during a memory task (de Cates et al., 2021) and decreases default mode network (DMN) functional connectivity (de Cates et al., 2023). In contrast, prucalopride does not show significant effects on emotional processing that mirror those seen with conventional antidepressants in healthy volunteers (Lucas et al., 2005; Lucas et al., 2007; Mendez-David et al., 2014).

Together this evidence positions 5-HT_4_R agonism as a promising approach for treating cognitive dysfunction in depression. The current study aimed to investigate the cognitive effects of prucalopride in individuals with a history of recurrent depression, currently not meeting ICD-10 criteria for depression for at least six months and free of psychotropic medication. The study extends previous work by testing a higher dose of prucalopride (2mg) and by including a broader battery of cognitive tests to further characterise the effects of 5-HT_4_R agonism on attention and executive function, as well as memory. We hypothesised that prucalopride would exert a broad pro-cognitive effect in this population.

## Methods

### Participants

Participants between 18 and 40 years old and of either sex were recruited to the study. They were screened to exclude those with contraindications to serotonergic medication. All participants were currently free of significant medical or psychiatric disorder but had a history of at least two episodes meeting DSM-5 criteria for major depressive disorder, as determined using the Structured Clinical Interview for DSM-5 (SCID). Other criteria for inclusion were: English language fluency; at least six months recovery since most recent depressive episode; current PHQ-9 score of less than 10; no current use of any medication except contraception; no medical concerns related to use of prucalopride. Full exclusion criteria are detailed in Supplementary Table 1.

The study was approved by the University of Oxford Central University Research Ethics Committee (CUREC, R77135/RE005). The protocol was pre-registered with clinicaltrials.gov (NCT05220228). Participants gave written informed consent. Full details of all methods are in the Supplementary Material.

### Design and randomisation

#### Overview of design and randomisation

Participants were randomised to one of two groups: prucalopride (2 days x 1mg followed by 5-8 days x 2mg), or placebo (lactose tablets for 7-10 days), in a double-blind, randomised, between-groups design. Randomisation was stratified by gender and was conducted using an online randomisation tool with block size of 4 (https://www.sealedenvelope.com/simple-randomiser/v1/lists).

Prucalopride and placebo tablets were encapsulated using a standardised operating procedure to ensure that they appeared identical. Participants received two bottles to take consecutively: bottle one had two capsules of 1mg prucalopride or placebo (for first two days of administration); bottle two had eight capsules of 2mg prucalopride or placebo. The full licensed dose of prucalopride (2mg) was used for at least five days to achieve steady state (terminal half-life approximately 24 h). Daily reminders optimised compliance.

Our previous translational studies of prucalopride in healthy volunteers employed a daily dose of 1mg which was well-tolerated. However, 2mg daily is the licensed dose for prucalopride and uncertainty exists regarding the dose required for full effect at brain 5-HT_4_ receptors. As previous studies in healthy volunteers suggested that a single dose of prucalopride has pro-cognitive effects on declarative memory with effect sizes ranging from d=0.5-0.9 (S. Murphy, Wright, Browning, Cowen, & Harmer, 2020a), in this study (with a higher dose and longer duration), we conservatively estimated d=0.5-0.7, and thus calculated 23 participants per group would be needed for power of 85-90% and alpha of 0.05.

#### Study visits

Two study visits (baseline and follow up testing after at least seven days of prucalopride / placebo) took place at the Neurosciences Building, Department of Psychiatry, Warneford Hospital following initial screening. At the point of post-intervention testing, prucalopride blood levels at 2mg daily would be expected to be at a steady state.

### Questionnaire measures

Participants completed the following self-report questionnaires to obtain baseline measures of mood, affect, cognitive difficulties, anhedonia and trait anxiety: PHQ-9; Positive and Negative Affect Scale (PANAS) (Watson, Clark, & Tellegen, 1988); Perceived Deficits Questionnaire (PDQ20) (Sullivan, Edgley, & Dehoux, 1990); Snaith–Hamilton Pleasure Scale (SHAPS) (Snaith et al., 1995); Spielberger State-Trait Anxiety Inventory, Trait Version (STAI-T) (Spielberger, Gorssuch, Lushene, Vagg, & Jacobs, 1983). Post intervention, at the start of the research testing visit, participants’ state anxiety and affect were measured using the Spielberger State-Trait Anxiety Inventory, State Version (STAI-S) (Spielberger et al., 1983) and the PANAS. Binary data regarding the most commonly reported side effects of prucalopride (Frampton, 2009) were measured both pre- and post-intervention to give a measure of side effects across the course of the study. At the end of the study, participants guessed their drug allocation with a forced-choice question.

### Cognitive tasks

We evaluated participant performance across a range of “cold” cognitive tasks that did not contain emotionally-valenced stimuli (the auditory verbal learning and memory task testing declarative memory (AVLT), a working memory task (N-back), and tests of executive functioning (including attention and processing speed: TMT, DSST)). We also used “hot” cognitive tasks that included a component of processing relating to emotionally-valenced information. These included the Emotional Go/No-Go (E-GNG) task, the Emotional Test Battery (ETB), and the Facial Attentional Dot Probe Task (FDOT). Multiple versions were available for the AVLT, N-back, TMT, and E-GNG, enabling data collection at baseline and post-intervention; other tasks were conducted post-intervention only. Apart from for the AVLT (auditory stimuli and verbal responses) and DSST and TMT (paper-based), all task stimuli were presented on a computer screen and participants required to respond via button presses on a keyboard. Full details regarding all tasks are given in Supplementary Material, and each is outlined briefly here. Non-emotional tasks were pre-determined as primary and emotional tasks as secondary for analysis purposes (see clinicaltrials.gov NCT05220228).

#### “Cold” cognitive tasks

##### Auditory Verbal Learning Task

In the Auditory Verbal Learning Task (AVLT) (see Figure 1(A)), participants’ verbal learning and memory was assessed using a multi-trial word recall paradigm. 15 concrete nouns (List A) were read aloud across five consecutive learning trials. After each presentation, participants were asked to immediately recall as many words as possible. Following the fifth trial, an interference list comprising 15 unrelated words (List B) was presented, with immediate recall subsequently assessed. Participants were then asked to recall the original List A words after a short delay (short-delay recall) and again following a longer delay of approximately 20 minutes (long-delay recall). Outcome measures included the total number of correct words recalled, repetitions (correct words recalled more than once in the same acquisition trial), and intrusions (incorrect words not present in the list).

**Figure 1.**
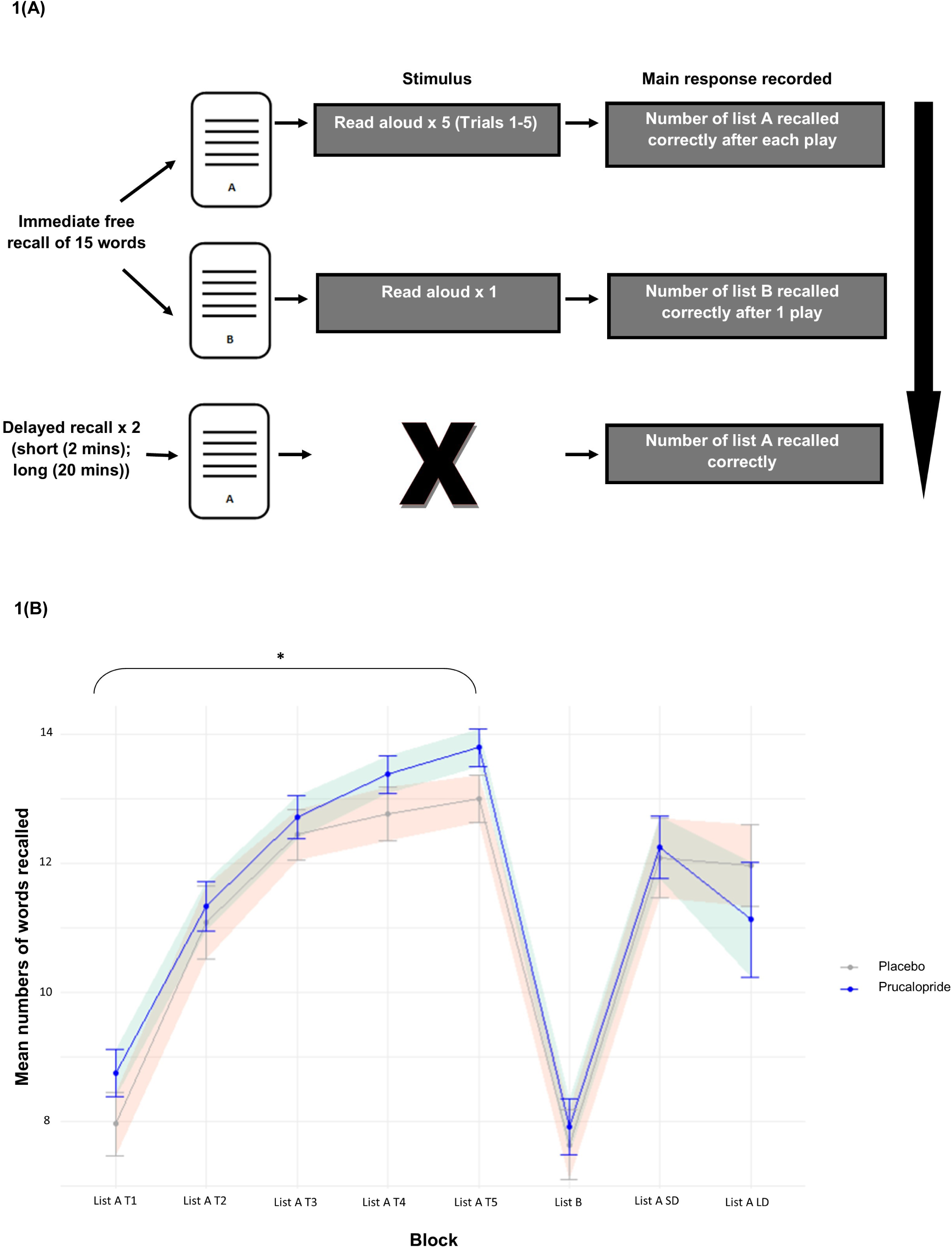
Mean number of words recalled across the Auditory Verbal Learning Task (AVLT) comparing the prucalopride and placebo groups. (A) Participants were read 15 concrete nouns from List A at a rate of one word per second. Participants were asked to immediately verbally recall as many items as they could, in any order. This was repeated a further four times, comprising five acquisition trials. Participants were then read a second set of 15 nouns (List B), and asked to recall words from this second list only. Immediately following List B, participants were asked to recall List A (short-delay), and once more after a delay of approximately 20 minutes, during which another task was completed (long-delay); (B) Error bars and shaded area indicate standard error of the mean. * represents statistical significance at p=0.05. Graphs with baseline data included are in Supplementary Material.

##### N-Back

In the N-back task (see Figure 2(A)), participants were required to indicate whether a visually presented symbol matched the one presented “n” trials earlier, where “n” corresponded to zero (0-back), one (1-back), two (2-back), or three (3-back). This task incrementally increased working memory demands across conditions. Performance was assessed based on reaction time and accuracy at each level of difficulty.

**Figure 2.**
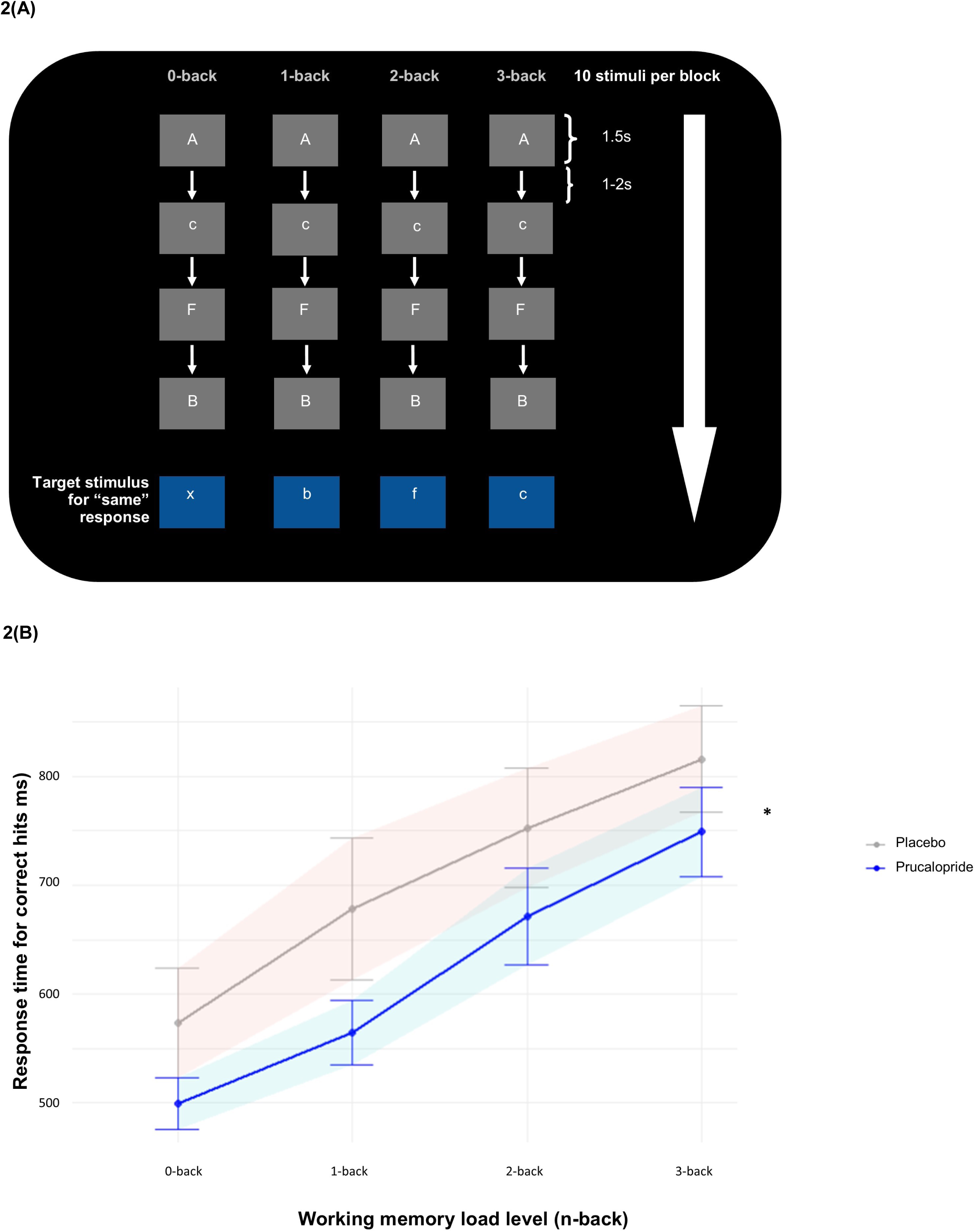
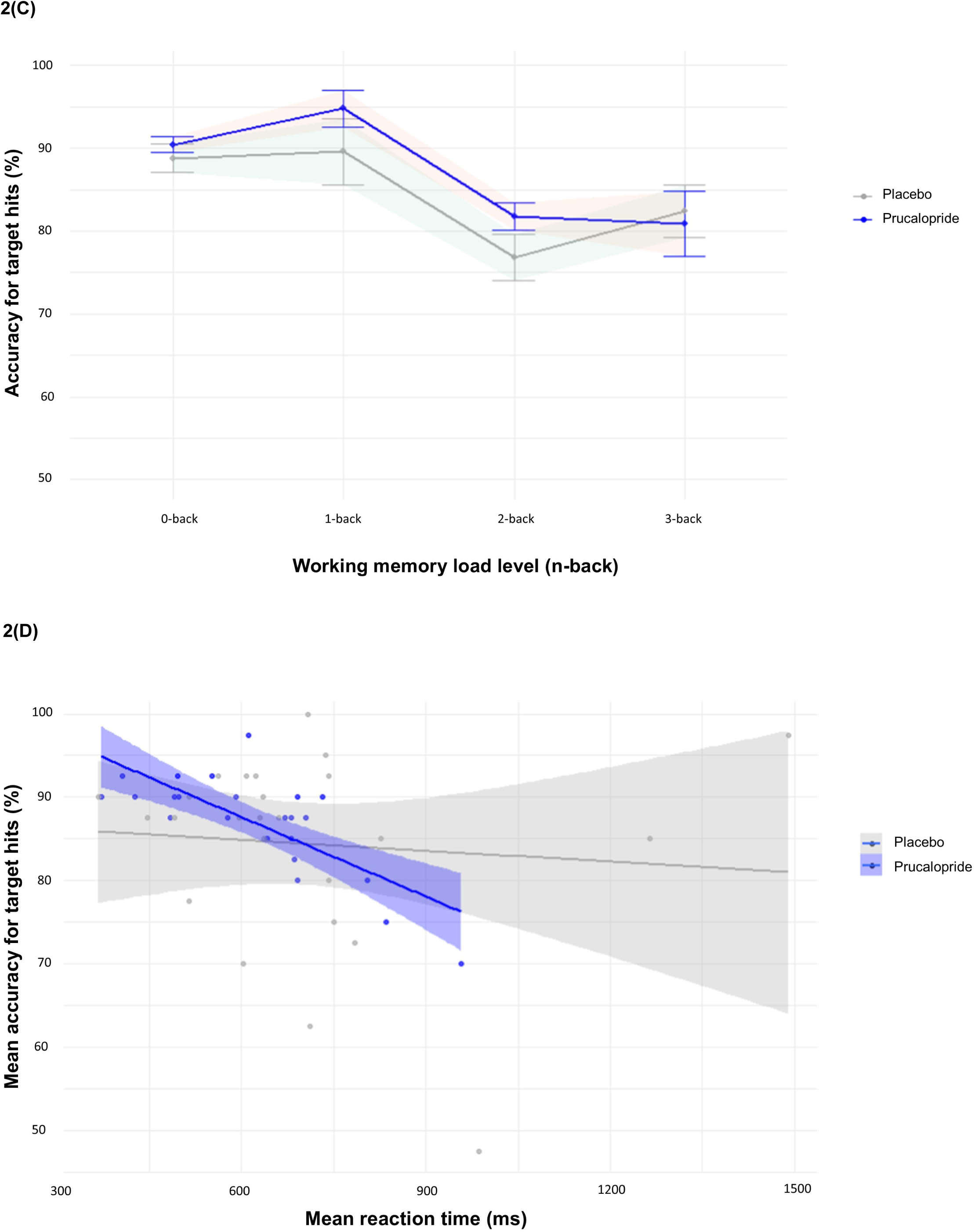
Results for the N-back task according to working memory load in the prucalopride and placebo group. (A) Verbal n-back task exemplar for conditions of 0-back, 1-back, 2-back and 3-back. Before each block of 10 stimuli per condition, participants were given specific instructions (e.g. “Press the spacebar if you see the same letter that appeared 1 letter ago” [1-back]). Each level of back was repeated four times (16 blocks total)]. For 0-back, participants were asked to select “same” only when the letter “X” appeared. For 1-back/2-back/3-back, participants were asked to select “same” if the current letter was the same as the one/two/three before; (B) Mean response time (ms) for correct hits; (C) Mean accuracy (%) for correct hits; (D) Mean accuracy compared to mean response time for correct hits. Error bars indicate standard error of the mean. Shaded area indicates 95% confidence interval. * represents statistical significance at p=0.05

##### Digit-Symbol Substitution Task

In the Digit-Symbol Substitution Task (DSST), participants were required to matching symbols to corresponding letters using a reference key. Performance was evaluated by recording the total number of correct symbols entries and the time taken to complete the task, providing a measure of processing speed and cognitive efficiency.

##### Trail-Making Task

The Trail-Making Task (TMT) assessed processing speed, attention and cognitive flexibility. It consisted of two parts: Part A required participants to connect consecutively numbered circles (e.g. 1-2-3), while Part B involved connecting circles in an alternating sequence of numbers and letters (e.g. 1-A-2-B). Performance was evaluated based on completion time and accuracy for each part.

Additionally the difference in completion time between Part B and Part A (B-A) was calculated as an index of executive functioning, specifically set shifting ability.

#### “Hot” cognitive tasks

##### Emotional Test Battery

The Emotional Test Battery (ETB) included four tasks assessing emotional processing. In the Facial Expression Recognition Task (FERT) (see Figure 3(A)), participants identified facial expressions (angry, disgusted, fearful, happy, sad, and surprised) presented briefly (500ms) on a computer screen.

**Figure 3.**
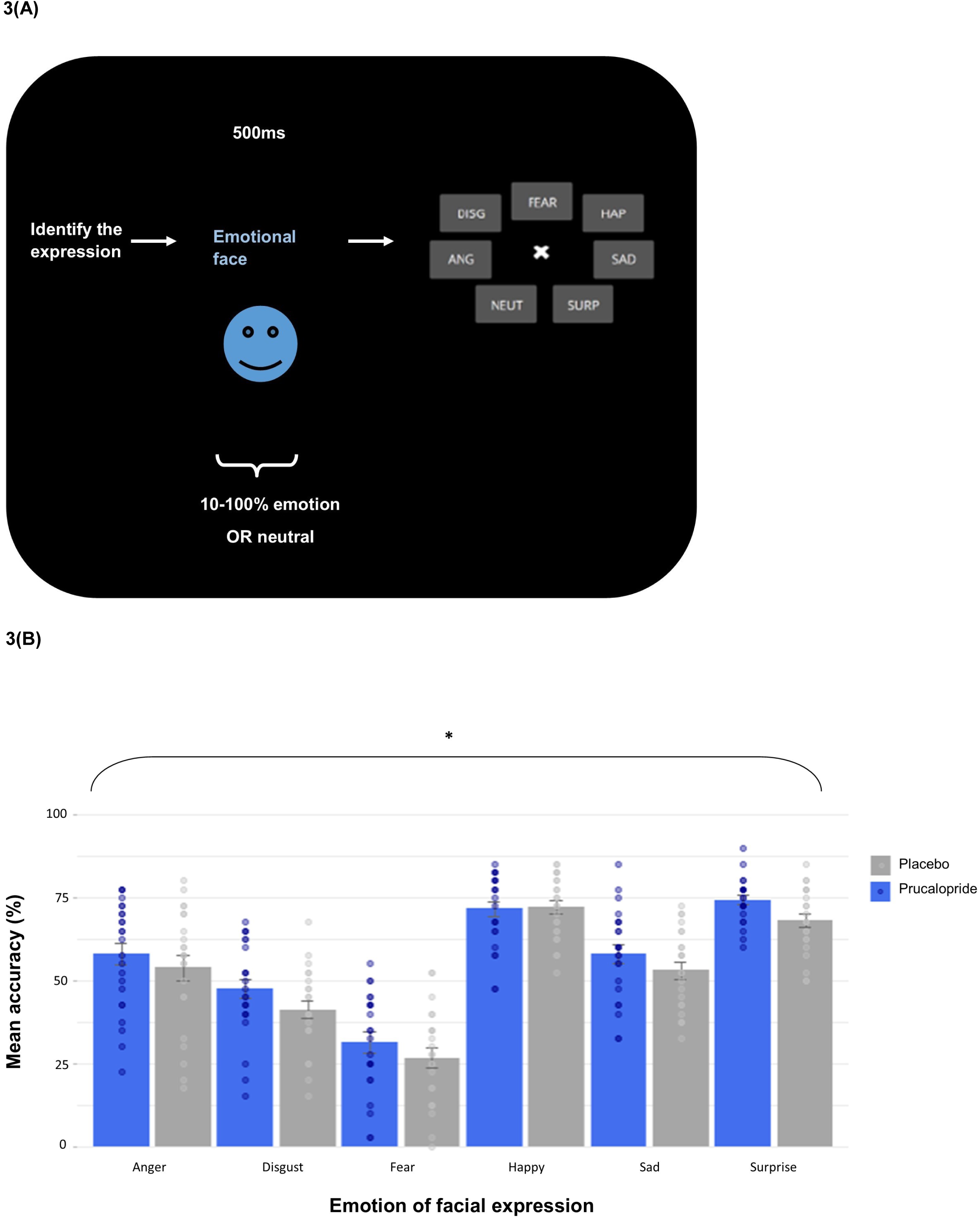
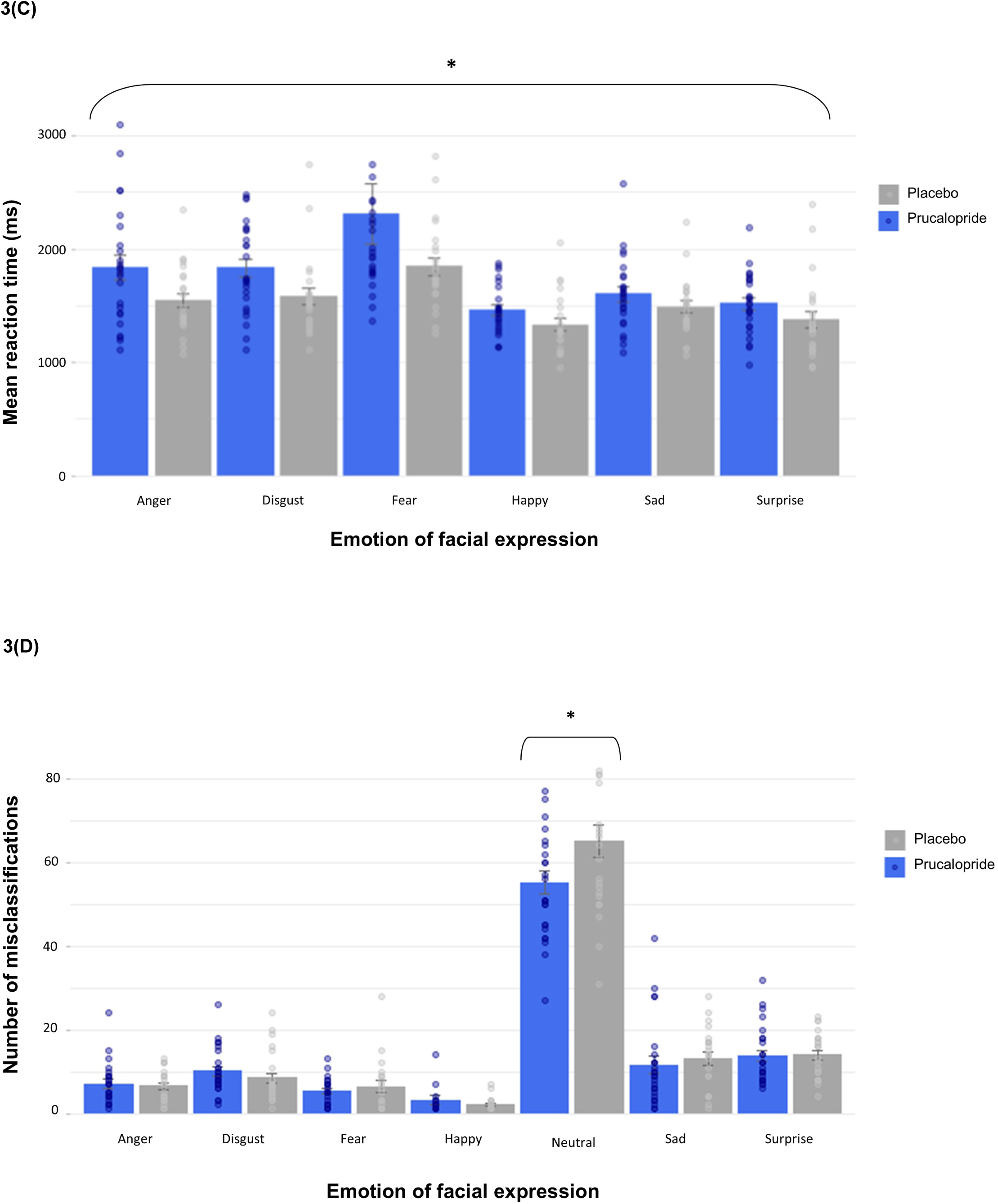
Mean accuracy and response time for correct responses and misclassifications on the Facial Expression Recognition Task (FERT) showing prucalopride and placebo group responses. (A) 250 randomised face images (emotions (anger, disgust, fear, sadness, surprise, and happiness) and neutral) were shown for 500ms in varying intensities in 10% gradations ranging from 0% (neutral) to 100% (full emotion). Participants were asked to identify the emotion presented; (B) Mean accuracy (%); (C) Mean reaction time (ms); (D) Mean number of misclassifications. Error bars indicate standard error of the mean. * represents statistical significance at p=0.05

Expressions were displayed at varying intensities, ranging from neutral to 100% full emotion in 10% increments. Accuracy to identify the correct emotion, reaction time and misclassification rates were recorded. In the Emotional Categorisation Task (ECAT), participants indicated whether they would like or dislike being described as positive and negative personality descriptor words. Classification accuracy and reaction times were measured. In the Emotional Recall Task (EREC), participants completed an unexpected free recall of the words previously presented in the ECAT, with the number of correctly recalled positive and negative words, as well as false alarms, recorded. Finally, in the Emotional Recognition Task (EMEM), participants identified whether a series of words presented on the screen were familiar (previously seen in the ECAT) or novel. Accuracy and response time were measured.

##### Faces Dot Probe Task

The Faces Dot Probe Task (FDOT) measures attentional vigilance to fear and happy faces, as previously described (S. E. Murphy, Yiend, Lester, Cowen, & Harmer, 2009). Participants were shown pairs of faces – one emotional (fearful or happy) and one neutral – positioned at the top and bottom of the screen. These were then replaced by a pair of dots (probe) aligned either vertically or horizontally. Participants were instructed to indicate the orientation of the dots by pressing a labelled key. On half of the trials (unmasked condition), the face pair was presented for 100ms and immediately followed by the probe. On the other half of the trials (masked condition), the sequence of events was exactly the same except the face pair was only presented very briefly (16ms) and followed by a mask (constructed from a jumbled face), which was displayed for 84ms. Attentional vigilance towards the emotional stimuli was assessed by calculating vigilance scores. These were derived by subtracting reaction times on incongruent trials (i.e. where the probe appeared in the location of the neutral expression) from congruent trials (i.e. where the probe appeared in the location of the emotional face), after reaction times for incorrect trials were removed. Positive vigilance scores indicated a bias towards emotional stimuli.

##### Emotional Go/No-Go Task

This task measured behavioural inhibition in the presences of affective interference, using happy, fearful and control (neutral/scrambled) images as distractors, as previously described(Colwell et al., 2024). Participants were instructed to respond (Go) or withhold responses (No-Go) based on shifting task rules (e.g. “Do not press the button if the image is yellow”) while simultaneously being presented with an emotional distractors (a fearful or happy face) or a control image. These task rules shifted across blocks. Emotional faces were irrelevant to the task rules but served as distractors to introduce emotional interference. Outcome measures included (1) accuracy on No-Go trials, reflecting response inhibition; (2) accuracy on Go trials; and (3) response times on Go trials, providing an index of impulsivity and processing speed.

### Data analysis

Behavioural data and questionnaires were analysed and graphs produced using RStudio (version 4.3.3). Two-sided t-tests, chi-squared tests, and analysis of variance (ANOVA) were used as appropriate for demographic and questionnaire analyses. Levene’s test (t-tests) and the Greenhouse–Geisser procedure (ANCOVAs/ANOVAs) were used as required. A repeated-measures analysis of co-variance (ANCOVA) was used as pre-specified to analyse group differences in behavioural performance on tasks suitable for repeated testing, with baseline data for the task used as the covariate (AVLT, N-back, DSST, EGNG) and estimated marginal means for planned comparisons after ANOVAs. For the AVLT, mean of list A scores across the first five blocks at baseline and task version order were included as covariates. For the Emotional Go/No-Go (EGNG), we also conducted drift diffusion modelling. A p-value less than 0.05 was used to denote statistical significance. Partial eta squared and Cohen’s D are reported as appropriate as measures of effect size. Sensitivity analyses involved the following included as covariates: baseline mood (PHQ9) and subjective cognition scores (PDQ20) (all tasks); age and years of education (executive functioning tasks (DSST, TMT)), native language (verbal processing tasks (EREC, ECAT, EMEM)). All sensitivity analyses are in Supplementary Material, and are reported where they differ from main analyses.

## Results

### Participants

In total, 50 participants were randomised into the study across two groups (placebo and prucalopride groups (25:25)) between 3^rd^ March 2022 and 23^rd^ October 2023 (see Supplementary Figure 1). Seven additional participants were recruited but replaced due to active Covid infection during the study or ineligibility determined after randomisation. One participant (prucalopride group) was excluded prior to unblinding for data quality issues, resulting in 49 participants for analysis (25:24; placebo:prucalopride). Analysis occurred in originally assigned groups. Data was not collected for the Emotional Test Battery tasks for one participant allocated to the prucalopride group as they had recently undertaken this during another study. Participants received the intervention for a similar period across groups [mean (SD): placebo 7.08 (0.28) days, prucalopride 7.25 (0.61) days].

At baseline, the groups were well matched in terms of demographics (i.e. age, sex, BMI, ethnicity, first language, years of education, handedness, and substance use; see Table 1), and severity of subjective cognitive problems (PDQ (Sullivan et al., 1990)), affect (PANAS (Watson et al., 1988)) and trait anxiety (STAI-T (Spielberger et al., 1983)). Mean PHQ-9 score at baseline indicated low depressive symptomatology (consistent with remitted depression) [Mean (SD): Placebo 3.08 (2.71); prucalopride 2.38 (2.24)], and was similar across the two groups.

**Table 1:** Baseline demographics, mood and affect symptoms for the Progress study.

| Variable (measure) | Measurement |  | Placebo<br>N=25 | Prucalopride<br>N=24 |
| --- | --- | --- | --- | --- |
| **Sex** | % | *Male* | 36.0 | 29.0 |
|  |  | *Female* | 64.0 | 71.0 |
| **Age** | *Mean in years (SD; range)* |  | 30.2 (9.95) | 28.0 (7.16) |
| **Years of education** | *Mean (SD)* |  | 17.6 (3.04) | 17.8 (2.63) |
| **First language** | % | *English* | 72.0 | 91.7 |
|  |  | *Not English* | 28.0 | 8.3 |
| **Ethnicity** | % | *White* | 56 | 50 |
|  |  | *Asian* | 28 | 16.7 |
|  |  | *Black* | 4 | 8.3 |
|  |  | *Other / Mixed* | 12 | 25 |
| **Current smokers** | % |  | 12.0 | 16.7 |
| **Alcohol** | *Mean units / week (SD)* |  | 2.53 (3.43) | 2.66 (3.39) |
| **Caffeine** | *Mean cups / day (SD)* |  | 1.53 (1.35) | 1.30 (1.10) |
| **Body mass index** | *Mean (SD)* |  | 24.7 (3.63) | 23.4 (3.25) |
| **Depression PHQ-9** | *Mean (SD)* |  | 3.08 (2.71) | 2.38 (2.24) |
| **Subjective cognition (PDQ-20)** | *Mean (SD)* |  | 25.3 (11.8) | 19.4 (11.7) |
| **Anhedonia (SHAPS)** | *Mean (SD)* |  | 0.60 (1.63) | 0.50 (0.83) |
| **Anxiety (STAI-T)** | *Mean (SD)* |  | 33.6 (8.7) | 38.0 (10.1) |
| **Affect (PANAS)** | *Mean (SD)* | *Positive* | 31.3 (6.57) | 33.8 (7.62) |
|  |  | *Negative* | 12.7 (3.12) | 11.8 (2.26) |

### Tolerability of prucalopride during the interventional period

Prucalopride was well tolerated compared to placebo administration. Binary ratings of side effects (present/non present during the last week) were analysed using 2-way ANOVAs with time point (pre-intervention, post-intervention) and group as factors. There were no significant time*group interaction differences in reports of total side effects [F(1,47)=0.12, p=0.73, np^2^<0.01], or headache, abdominal pain, nausea, diarrhoea, dizziness, vomiting, flatulence, or gastrointestinal sounds when analysed individually (all ps > 0.3, see Supplementary Table 2). There was a group*time interaction for the decreased appetite item, which reflected significantly higher scores in the prucalopride group post-intervention [F(1,47)=5.42, p=0.024; placebo participants endorsing at baseline=12% and follow-up=4%; prucalopride participants endorsing at baseline=12% and follow-up=24%]. Further detail is in Supplementary Material.

Randomisation guesses (data missing for one placebo participant) suggested that most placebo participants guessed their allocation correctly whereas prucalopride participants were at around chance levels [correct guess: placebo 83.0% (20/24), prucalopride 58.3% (14/24); ꭕ^2^= 4.61, p=0.10].

### Subjective anxiety, affect, and mood

There were no significant differences between the two groups in state anxiety and negative affect at the start of the post-intervention research visit [STAIS-S (mean (SD): Placebo: 30.7 (6.65) Prucalopride 31.7 (9.90); t(40.1)=-0.37, p=0.71); PANAS-N (mean (SD): Placebo: 12.32 (2.93) Prucalopride 12.5 (4.11); t(41.4)=-0.18, p=0.86)]. However, in the prucalopride group there was a borderline significant difference between the groups in terms of positive affect at the post-intervention research visit, with higher scores in the prucalopride group compared with the placebo group [PANAS-P mean (SD): Placebo 29.1 (6.98); Prucalopride 33.2 (7.35); t(46.6)=-1.99, p=0.05, Cohen’s d = -0.57 (−1.14, 0.00)], which was not wholly explained by PANAS scores at baseline (see Table 1)].

Subjective mood was not significantly different post-intervention compared to baseline across groups [mean (SD): baseline PHQ9 placebo 3.08 (2.71) prucalopride 2.38 (2.24); post-intervention QIDS-C placebo 4.16 (2.61) prucalopride 3.58 (2.50); F(1,94)=1.67, p=0.20], see Supplementary Figure 2 using equivalence tables from (Palmer, Ker, Rentería, Carmody, & Rush, 2024).

### Does 5-HT4 receptor agonism affect non-emotional “cold” cognition in those with remitted depression?

#### Declarative memory (Auditory verbal learning task (AVLT))

In both groups participants’ recall of words from List A improved across the five acquisition blocks (see Figure 1(B) and Supplementary Figure 3(A and B)). Participants in the prucalopride group were on average more accurate than those in the placebo group in their recall across these five acquisition blocks [F(1,226)=8.12, p=0.005, np2=0.18 (0.07-0.33)]. There was no statistically significant group*block interaction [F(4,226)=0.45, p=0.77, np2=0.05 (0.00-0.08)], indicating that recall was improved on average across all the task blocks.

There was no difference between the prucalopride and placebo participants in terms of the number of words accurately recalled after a short delay [F(1,42)=0.016, p=0.90] or a long delay [F(1,42)=0.96, p=0.33], or for intrusions, repetitions or recall of a separate set of words (List B) (all *p*s > 0.3, see Supplementary Figure 3(C)).

#### Verbal working memory (N-back)

The prucalopride group were significantly quicker on average across task loads (0- to 3-back) [F(1,179)=8.56, p=0.004, np2=0.03 (0.00-0.19), see Figure 2(B)]. There was no group*condition interaction (F(3,179)=1.51, p=0.21, np2=0.04. The prucalopride group tended to make correct choices more quickly although the group*reaction time interaction fell short of significance [group*mean reaction time: F(1,44)=3.17, p=0.082, np2=0.07], see Figure 2(D) and Supplementary Table 5. This was also reflected by the relationship between d’ and group (t(−1.68, 45.53), p=0.099, see Supplementary Figure 4). Findings were unchanged in sensitivity analyses, although when we excluded a placebo participant who was an outlier for reaction time (reduced speed) but not for accuracy the main effect of group for reaction time became borderline [F(1,175)=3.71, p=0.05, np2=0.01].

There was no main effect of group in terms of accuracy for recalled targets [F(1,179)=2.12, p=0.15, np2=0.01 (0.00-0.06)), although there was a trend for increased accuracy at the 1- and 2-back level [See Figure 2(C); 0-back EMM: -0.16 ± 0.40, *p* = 0.68, d=-0.12 (−0.69-0.45); 1-back EMM: - 0.52 ± 0.40, *p* = 0.19, d=-0.38 (−0.95-0.19); 2-back EMM: -0.49 ± 0.40, *p* = 0.21, d=-0.36 (−0.93-0.21); 3-back EMM: 0.15 ± 0.40, *p* = 0.70, d=0.11 (−0.46-0.68)]. Data from 1 participant (prucalopride) was missing for the N-back.

#### Tasks assessing executive function

There were no statistically significant group effects for performance for the Digit Symbol Substitution task [F(3,45)=-0.88, p=0.38] or the Trail Making Task [t(44.0)=0.007, p=0.99].

### Is the processing of emotions (“hot” cognition) affected by 5-HT4 receptor agonism in previous depression?

#### Facial expression recognition task (FERT)

Participants in the prucalopride group were more accurate than placebo participants across all emotions [F(1,270)=9.99, p<0.002, np2=0.02 (0.00-0.24)]. However, there was no interaction between group and emotion [F(5,270)=0.44, p=0.82], see Figure 3(B). Participants in the prucalopride group were also significantly slower than those in the placebo group across all emotions [F(1,269)=13.5, p<0.001, np=0.05 (0.02-1.00)]; with no group*emotion interaction [F(5,269)=0.65, p=0.66], see Figure 3(C).

Consistent with the overall effect of group on accuracy, the prucalopride group made fewer misclassifications of other faces as neutral [F(1,46)=4.47, p=0.04, np2=0.09 (0.00-1.00)], see Figure 3(D).

#### Faces dot probe task (FDOT)

The prucalopride group were significantly slower than the placebo participants across all trials [F(1,733)=9.06, p=0.0027, np2 = 0.01 (0.00-1.00), Placebo mean (SD) 0.55s (0.08), Prucalopride mean (SD) 0.57s (0.06), see Figure S7(A)]. There was no statistically significant group level effect in terms of attentional vigilance [F(1,686)=0.28, p=0.60, see Figure S7(B)]. There was a group*mask*emotion interaction [F(1,686)=4.19, p=0.04, np2 = 0.006 (0.00-0.10)]; however, this was no longer significant when examining group*emotion interactions for the masked and unmasked conditions separately [masked: F(1,344)=2.42, p=0.12; unmasked: F(1,342)=1.81, p=0.18]; see Figure S7(C) and Supplementary Table 11. Results were unchanged when we calculated proportional difference scores instead of vigilance to take account of the group-level difference in reaction time (see Supplementary Material).

#### Emotional Go/No-Go Task

5-HT_4_R agonism was not associated with a change in response inhibition (measured by mean percentage of accurately withheld responses to no-go trials) (ANCOVA main effect of group: F[1,46]=0.065, *p*=0.799, np2 < 0.01; Placebo mean (SD) 34.8 (8.4); Prucalopride mean (SD) 34.4 (7.9); all conditions EMM = 0.31 ± 1.84, *p* = 0.87 (Figure S8(A)), a group by set shift interaction for accuracy of withheld presses (ANCOVA: F[1,340]=0.107, *p*=0.74), or a group effect for go trial accuracy (ANCOVA main effect of group: F[1,46]=0.004, *p*=0.95) when considering either the control condition or affective interference (Figure S8(B)).

Prucalopride allocation also did not affect choice impulsivity, indicated by no change in reaction time to choice for go trials, across all task conditions (ANCOVA main effect of group: F[1,46] = 0.20, p = 0.63; ηp2 < 0.01) (see Figure S8(C)).

Signal detection theory analyses suggested that prucalopride allocation did not influence decision bias across task conditions (log criterion c; ANCOVA main effect of group: F[1,46] = 0.96, *p* = 0.33, η ^2^ < 0.01 [0.00, 0.00]; all conditions EMM = -0.015 ± 0.03, *p* = 0.58) (see Figure S8(D)); no other significant differences were evident between groups in drift diffusion modelling (see Supplementary Table 10 and Figure S8(E)).

Controlling for baseline mood and subjective cognition did not affect results (see Supplementary Material). When we excluded two participants (1 prucalopride, 1 placebo) each with trials where reaction times were outside optimisation parameters (i.e. too short indicating lack of engagement), results were similar apart from log criterion analyses (log criterion c), which became significant (see Supplementary Material).

#### Emotional Test Battery

There were no statistically significant group effects for performance or reaction time on the Emotional Categorisation Task (ECAT), the Emotional Recall Task (EREC) and the Emotional Recognition Memory Task (EMEM): all ps>0.3. This included when sensitivity analyses were performed controlling for baseline mood and subjective cognition scores (see Supplementary Material).

## Discussion

In this experimental medicine study of participants with a history of depression, seven to ten days of prucalopride administration, compared to placebo, was associated with improved performance on several cognitive measures. Specifically, prucalopride improved immediate recall on an auditory verbal memory task (AVLT), increased accuracy in facial expression recognition (FERT) regardless of emotional valence, and led to faster responses on a complex working memory task (N-back) with a trend toward improved accuracy.

These observed pro-cognitive effects of prucalopride (2mg) are consistent with previous translational work in healthy volunteers using a 1mg dose. In earlier studies, prucalopride improved multiple cognitive domains, including declarative memory, reward learning, emotional memory, and face processing (de Cates et al., 2021; de Cates et al., 2022; S. Murphy et al., 2020a). Our findings extend these effects to a remitted depressed sample and replicate improvement in immediate recall. Importantly, we demonstrate additional benefits with a broader profile of pro-cognitive effects, including on tasks of working memory.

On the N-back task, the prucalopride group responded faster across difficulty levels, with a trend toward improved accuracy. This contrasts with previous studies using a single 1 mg dose, which did not detect significant effects of prucalopride on working memory (S. Murphy et al., 2020a). In the current study, participants reached steady-state dosing (≥ 5 days) of 2mg, potentially enhancing observable effects in this cognitive domain. Our findings are also consistent with other work showing that 5-HT_4_R agonism is associated with faster reaction times without accuracy impairments in people with depression (de Cates et al., 2025).

Interestingly, reaction time patterns varied across tasks. Compared to placebo participants, the prucalopride group was slower on tasks involving emotional processing (FERT, FDOT), potentially reflecting more cautious and accurate responding. Conversely, as discussed, the prucalopride group was significantly faster on the N-back task, which reflects non-affective working memory.

There was also limited evidence that 5-HT_4_R agonism influenced emotionally-valenced cognitive performance; improvements on the FERT were not emotion-specific, and no significant effects on accuracy were observed for the Emotional Go/No-Go, FDOT, ECAT, EREC, or EMEM tasks. These limited effects of prucalopride on emotional cognition are consistent with previous work (de Cates et al., 2022) and with another recent 5-HT_4_R study from our groups (unpublished data). In this study, participants with current depression showed mood improvements following sub-acute administration with both an unlicensed 5-HT_4_R agonist (PF-04995274) and citalopram but only citalopram altered emotional processing biases – a key mechanism through which conventional antidepressants exert their effects (Harmer, Goodwin, & Cowen, 2009). This suggests that prucalopride’s mechanisms may be distinct from emotional bias modification.

Several neurobiological mechanisms may underlie the pro-cognitive effects of 5-HT_4_R agonism. First, 5-HT_4_Rs are densely expressed in the hippocampus, and rodent studies show improvements in hippocampal-dependent learning and memory following 5-HT4R activation (Lamirault & Simon, 2001; Marchetti et al., 2011; Mohler et al., 2007; Pfizer, 2011). Potential mechanisms include modulation of AMPA receptor-mediated glutamatergic transmission (promoting stress resilience) (Chen et al., 2020), increased acetylcholine release (Hagena & Manahan-Vaughan, 2017), and enhanced neurogenesis and dendritic growth (Lucas et al., 2007). Given the known hippocampal abnormalities in depression – including reduced hippocampal volume and disrupted functional connectivity with the prefrontal cortex and amygdala (Cao et al., 2012; Hao et al., 2020; Sheline, Liston, & McEwen, 2019; Cullen et al., 2014; Hamilton & Gotlib, 2008) – these mechanisms may have particular clinical relevance. Second, 5-HT_4_R agonism may exert effects on cognition and mood via the gut-brain axis. The vast majority of serotonin is produced in the gut, and peripheral serotonergic signalling may influence central cognitive flexibility (Shine et al., 2022). Notably, antagonism of 5-HT_4_Rs has been linked to pro-inflammatory microbiome profile, suggesting these receptors may also regulate gut-brain inflammation (Cui et al., 2023).

Given the prevalence of persistent cognitive problems after remission of depressive illness (Rock et al., 2014), and their association with relapse risk (Halahakoon et al., 2019), interventions that target and improve cognition could offer meaningful clinical benefits. Our findings were unchanged by including mood or subjective cognition scores at baseline into analytical models, suggesting that the potential pro-cognitive impact of 5-HT_4_R agonists may be relevant across mood states, including for individuals with subclinical or unrecognised cognitive impairments.

There are some limitations of the current study that should be considered. The study was underpowered for some tasks due to missing data (e.g. TMT sample was reduced from n=24 to n=22 in the prucalopride group). Although our sample included participants with varied histories of antidepressant use and number of previous episodes – supporting generalisability - it was predominately female, white and highly educated, and limited to adults under the age of 40. These factors may restrict the applicability of findings to broader, more diverse populations.

In conclusion, this is the first study to assess the cognitive effects of 5-HT_4_R agonism in a remitted depressed population. We replicated and extended prior evidence from healthy participants, demonstrating a pro-cognitive effect of prucalopride across verbal memory, face processing and working memory tasks. These effects appeared independent of emotional processing changes, highlighting a distinct neuropsychological profile compared with conventional antidepressants.

Future work should explore the therapeutic potential of 5-HT_4_R agonists in patients with depression and cognitive problems, investigate mechanisms of mood change outside of the emotional bias model, and further elucidate how 5-HT_4_ agonists may impact on the gut-brain axis. Given that cognitive deficits are an important feature of multiple serious mental illnesses, including schizophrenia and bipolar disorder, these findings may have broader transdiagnostic relevance.

### Financial support

ANdeC is currently funded by an NIHR Clinical Lectureship and also receives funding and support from the NIHR Mental Health Translational Research Collaboration (MH-TRC) Mental Health Mission and the NIHR Oxford Health Biomedical Research Centre. She has previously received funding from the Guarantors of Brain and a Wellcome Trust Clinical Doctoral Research Fellowship (216430/Z/19/Z).

This research was supported by the NIHR Oxford Health Biomedical Research Centre. The views expressed are those of the authors and not necessarily those of Wellcome, the NHS, the NIHR or the Department of Health. None of these bodies had a significant role in the design, collection and analysis of data, or decision to publish this article.

## Ethical standards

The authors assert that all procedures contributing to this work comply with the ethical standards of the relevant national and institutional committees on human experimentation and with the Helsinki Declaration of 1975, as revised in 2008.

## Data Availability

All data produced in the present study are available upon reasonable request to the authors

