## Supplementary Material for "Pro-cognitive effects of 5-HT4 receptor agonism in individuals with remitted depression"

### Supplementary Data

### Supplementary Methods

#### Power calculation

Previous studies examining the effect of prucalopride and other serotonergic drugs on neurocognitive outcomes, including learning / memory and emotional processing, indicate that an effect size of 0.8-0.9 may be expected [11, 13]. However, given relatively little is known about the effects of 5-HT_4_ receptor stimulation on cognition in humans, we took a conservative approach and calculated the samples size needed on the basis of a smaller estimated effect size (0.5-0.7). This indicated that 17 participants per group are needed to give 90% power to detect a significant difference between the two groups with an α of 5%. We therefore aimed to recruit 50 (25 in each group) as part of oversampling.

#### Recruitment

Participants were recruited via adverts emailed to and displayed in colleges and university departments and local community buildings, placed on local information websites (e.g. Daily Info, Oxford University Gazette), newspapers, local magazines, and on the lab webpage and Social Media (including Facebook). The adverts contained brief information about the inclusion criteria for the study, as well as contact details for the named researchers. We also collaborated with Lindus Health who specialise in recruiting participants for clinical trials.

After a potential volunteer contacted the research team, they were sent the Participant Information Sheet (PIS) via email. The participant was be given as much time as they need to decide whether they would like to take part, and invited to ask any questions that they have about the study. If the participant decided they would like to take part, they were invited for a screening visit.

#### Screening and consent

The screening visit consisted of two parts. Part 1 was conducted either remotely (as a videoconference or telephone call) or in person at the Department of Psychiatry, Warneford Hospital. Informed Consent was collected prior to the administration of any screening procedures. If Part 1 of the screening visit was held remotely, the researcher went through the informed consent form (ICF) point-by-point with the participant on the video call and completed a paper version of the ICF on their behalf.

The PIS was presented to the participants in written as well as in verbal form as part of the consent process. This detailed the exact practical demands of the study, written from the participant's perspective, in lay language (the nature of the study, what it will involve for the participants, the implications and constraints of the protocol, the known side effects and any risks involved in taking part, what will happen to the data collected). It was clearly stated that the participant was free to withdraw from the study at any time for any reason and with no obligation to give the reason for withdrawal. The participant was allowed as much time as they wish to consider the information, and the opportunity to question the Investigator, their General Practitioner (GP) or other independent parties. The participant signed and dated the latest approved version of the ICF before any study specific procedures were performed, with a counter signature of the investigator who took consent, and a copy given to the participant. If the first part of the screening occurred remotely, the participant later physically signed the ICF at the start of Part 2 of the screening visit, which always occurred in person. All of the named researchers were trained in taking informed consent.

#### Study visits

Study visits took place at the Neurosciences Building, Department of Psychiatry, Warneford Hospital. During the screening (baseline) visit, all participants underwent a medical evaluation including a review of medical history, current and past medication, weight, height and BMI measurement, urine drug screening, and a pregnancy test where relevant. Participants meeting all study criteria underwent baseline assessments (questionnaires and baseline cognitive assessments) and were given medication to take at home. During the period of drug / placebo administration, participants were advised not to drink alcohol and not to carry out activities requiring full alertness if they were aware of any impairment. A researcher also contacted them by phone on day two and day four to check that there were no concerns. Participants received a daily text message reminding them to take the study medication and had access to a 24h number in case of any concerns. Spare medication was given to all participants to enable completion of study visits up to and including day 10 to allow for scheduling difficulties or illness. Participants returned for a follow-up testing visit after taking at least seven days of prucalopride or placebo. At the point of post-intervention testing, 2mg prucalopride would be expected to be at a steady state.

#### Baseline assessments

Participants were invited to attend a Screening Visit (in 2 parts totalling approximately 2 hours). Part 1 of this was able to be conducted remotely using university videoconferencing software (Microsoft Teams) or a telephone call before attending Neurosciences in person, as detailed above.

Part 1:

Demographic information was collected:

- Age (years, months), years in full-time education

Part 1:

Participants' eligibility to take part was checked by recording the following:

- Number of cigarettes smoked per day, units of alcohol consumed per week, caffeinated drinks consumed per day
- Current and prior use of illicit substances
- Menstrual cycle (in order to avoid scheduling testing in the premenstrual week)
- SCID-V interview (Structured Clinical Interview for DSM-5) to probe for current or past psychiatric illness

Part 1&2:

To ensure it is safe to administer the drug to the participant, the following was recorded:

- Medical history (including family psychiatric history)
- Details of current and past medication, and any involvement in other research studies
- Details of contraception if female and child-bearing age, and confirmation that not breastfeeding
- Weight, height, and BMI
- Urine pregnancy test (female participants only)

Part 2:

The following baseline assessments were completed:

- Patient Health Questionnaire (PHQ-9) – self-report of depressive symptoms
- Perceived Deficits Questionnaire (PDQ-20) – a measure of subjective cognitive dysfunction
- Snaith-Hamilton Pleasure Scale (SHAPS) – a measure of anhedonia
- State and Trait Anxiety Inventory (STAI)
- Positive and Negative Affect Schedule (PANAS)
- Side effect questionnaire – to determine baseline of bodily symptoms/’side effects’
- Digit Symbol Substitution Test (DSST) – measure of executive function / attention / processing speed
- Emotional go/no-go – a measure of cognitive control
- N-back – a measure of working memory
- Auditory Verbal Learning Task (AVLT) – a measure of memory ability

Participants were asked to wait after part 2 while their eligibility to take part was assessed by a medical doctor involved in the study (maximum 30 minutes).

#### Randomisation

Eligible participants were randomised, using an online randomisation tool, to receive either seven-to-ten day prucalopride, or seven-to-ten day placebo administration. Randomisation occurred on the day of screening (Part 2 completion), or up to a maximum of four weeks after screening.

Randomisation was stratified for gender and blocked with a block size of 4. In order to maintain blinding, the drug or placebo were placed into containers labelled with each participant ID at the start of the study by a researcher not involved in the study, according to the randomisation schedule. The participants and study researchers were blinded to intervention until the end of the study. All assessments were administered by study researchers who were blind to group allocation.

Randomised participants were given seven days of the drug to take home, as well as full instructions of how and when to take them. There were two sets of tablets given to all participants, so that participants randomised to prucalopride received the lower 1mg dose for the first two days, and then if tolerated, the higher 2mg dose for the next five to eight days. The study was a double blind design thus neither the researchers nor the participant knew at this stage whether prucalopride or placebo was given.

During the week of drug/placebo administration, participants were advised not to drink alcohol and not to carry out activities requiring full alertness, such as driving, if they were aware of any impairment. During this week, a researcher phoned the participant on day 2 and day 4 to check that there were no concerns. There was a further contact on day 7, either in person for testing or remotely if testing is delayed. On day 2, if the participant was feeling well, they were advised by the researcher to continue onto the second set of tablets. Concerns at any check-in contact were discussed with a medically-qualified member of the study team. Additionally, participants received a text message every day reminding them to take the study medication. They also were given the 24-hour contact phone number of a member of the study team and encouraged to get in contact if they had any concerns or queries during the study week, or if their medication or health status changed.

#### Research visit

Research Visit (approximately 3 hours) took place on day 7-10 of prucalopride/placebo administration in the Neurosciences Building, Warneford Hospital. This visit involved administration of behavioural tasks measuring emotional and non-emotional cognitive processing.

Participants completed the following questionnaires before the behavioural tasks:

- State Anxiety Inventory (STAI-S)
- Positive and Negative Affect Schedule (PANAS)
- Side effect questionnaire

The non-emotional processing behavioural tasks included tests of attention / executive functioning, processing speed, cognitive control, memory, and a reward task: DSST, TMT, emotional go/no-go, N-back, AVLT. The FDOT and emotional processing tasks were from the Emotional Test Battery (ETB), which has previously been found to be sensitive to the effects of antidepressants. These are detailed in the Tasks section.

With the exception of the DSST and TMT (paper-based), and AVLT (auditory-based), all stimuli were presented on a computer screen and participants required to respond via button presses on a keyboard. Tasks performed at baseline (screening) and repeated at the research visit used different versions to minimise learning effects.

At the end of the research visit, a member of the study team asked participants to try to guess which treatment they received, as a measure of blinding success.

#### Tasks

Auditory Verbal Learning Task (AVLT): The AVLT measures declarative verbal memory. Participants were read 15 concrete nouns at a rate of one word per second (List A, e.g. Drum, Curtain). Participants were asked to immediately verbally recall as many items as they could, in any order. This was repeated a further four times, comprising five acquisition trials. Participants were then read a second set of 15 nouns (List B), and asked to recall words from this second list only. Immediately following List B, participants were asked to recall List A (short-delay), and once more after a delay of approximately 20 minutes, during which another task was completed (long-delay). Number of words correct, repetitions (correct words recalled more than once in the same acquisition trial), and intrusions (incorrect words not present in the list) were measured. We stratified for version across the groups. For analysis of the post-intervention AVLT scores, and accounted for version order and baseline AVLT list A mean score (mean score across the first five trials) as per standard practice. We ran supplementary analyses evaluating the effect of version order on the outcome of list A mean scores at the research visit to confirm its inclusion in the model (see Supplementary Results).

N-back: This letter variant of the N-back task measures working memory [8]. Participants were shown a series of letters, and asked to respond by clicking either the key “same” or “different” depending on the task condition. This task had four conditions. In the 0-back condition, participants were asked to select “same” when the letter “X” appeared, and different for all other letters. In the 1-back condition, participants were asked to compare each letter with the one before and select “same” if the current letter was the same as the one before. In the 2-back condition, participants were asked to compare between the current letter and the letter shown two places ago. Finally, in the 3-back condition, participants were asked to compare between the current letter and the letter shown three places ago. Each trial starts with a screen stating the condition, followed by a central fixation cross. The end of the trial is indicated by the appearance of a central fixation cross. Data analysis for this task involved looking at reaction time and accuracy.

Digit Symbol Substitution Test (DSST). This task measures processing speed, attention, and visual-motor coordination. Participants match symbols to corresponding numbers within a time limit. Both tests are sensitive to cognitive decline and are commonly used in clinical settings and research. Time to complete the task was measured.

Trail-Making Task (TMT): This task consists of two parts: Part A primarily assesses visual attention and processing speed, while Part B adds cognitive flexibility and executive function. Part A requires connecting numbered circles in sequence, while Part B alternates between numbers and letters (1-A-2-B-3-C, etc.). Time to complete each part was measured. The time to complete Part B minus Part A was then calculated.

Facial Expression Recognition Task (FERT): The FERT measured a participant’s ability to differentiate between 6 basic emotions (anger, disgust, fear, sadness, surprise, and happiness) and neutral, presented as images of facial expressions. Face images were selected and modified from the Karolinska Directed Emotional Faces (KDEF) set. The images were shown in varying intensities in 10% gradations ranging from 0% (neutral) to 100% (full emotion). A total of 250 (10 individuals contributing a total of 4 examples of each emotion at each intensity, plus a neutral expression for each individual) randomised images were presented on a computer monitor (500 ms per image). Each version of the task uses images created from four models and 60 images have been created for each model (six emotions at 10 levels of intensity). Over all blocks, each image is presented only once. Neutral faces images (0% emotion) are also presented in 10 trials, with two repeats of the neutral expression for two models, and three repeats of the neutral expression for the two other models. The order of images is randomised within and between blocks, with the same randomised order used for all runs of the task. The fixed randomisation ensures that there are no more than two contiguous trials of the same emotion (at any percentage), no more than two contiguous trials of the same model, no contiguous trials with the same model and same emotion, and balanced numbers of emotions across all blocks (10 of each, plus 2 or 3 neutral trials). Analysis of the data involved looking at accuracy scores, reaction time, and misclassifications. Accuracy scores comprised of total accuracy (total number of correct responses for each emotion). Misclassifications were defined as the number of responses when the participant chose a facial expression when it was actually an expression of a different category. For all accuracy and reaction time analyses, emotions were defined as the 6 emotions (anger, disgust, fear, happy, sad, surprise) and neutral was considered as baseline and not included in the analyses.

Emotional Categorisation (ECAT), Emotional Recall (EREC), and Emotional Memory (EMEM):

For the ECAT, participants were presented with 40 personality characteristic words on a computer screen for 500 ms. Words were selected to be positive or negative in nature (20 each), and were matched for length, frequency and meaningfulness: negative words included “domineering”, “untidy”, “hostile”; positive words included “cheerful”, “honest”, “optimistic”. The word lists were updated in 2021, following new data collection (based on Anderson, 1968; Raslescu et al, 2022). Each word is presented once over the course of the experiment. For each word, participants were asked to imagine overhearing someone describing them with that characteristic, and then determine if they would like or dislike to be described in that manner. There was a fixed 3000 ms intertrial-interval (response period) which follows stimulus offset during which the participant is required to press a key to identify whether that personality characteristic is one they would “like” (key press Q) or “dislike” (key press O) to be described as. We analysed the number of classifications, and response times for correct identification.

The EREC is a surprise recall task where participants were given 4 minutes to write down as many characteristic words as they could remember from the ECAT. All words were scored independently by two raters, with a group consensus for disagreements. Spelling mistakes did not result in a word being marked as incorrect. Slight variations were not marked as incorrect as long as these did not change the meaning of the word. Emotional biases in memory was measured as the relative recall of positive versus negative words.

In the EMEM, participants were presented with a series of personality characteristics as words on a computer screen. Half of the presented words were positive and negative words previously seen by participants in the ECAT, while the other half were novel distractor words. There were three blocks of words, separated by a rest period of length determined by the participant (participant controls progression to the next block). The first block contains 28 words and second and third block contain 27 words. Within each block the 20 positive and 20 negative personality characteristic words presented in the ECAT are again presented, along with 20 novel positive personality characteristic words and 20 novel negative personality characteristic words. The order of words presented was newly randomised with each run of the task. Novel words were matched with familiar words in terms of word length and ratings of frequency and meaningfulness. Participants were asked if they had previously seen the word (familiar) in the ECAT or not (novel). There was a fixed 3000 ms inter-trial-interval (response period) which followed stimulus offset. This task measured emotional biases in memory, and we analysed the accuracy and response time of the recall, and the valence of the recall (i.e. whether positive information was remembered better).

Facial Attentional Dot Probe Task (FDOT): Participants were shown a display where a pair of faces would appear (one on the top half of the screen and the other at the bottom half), followed by 2 dots that were either in vertical or horizontal orientation in the location of one of the faces. Participants were asked to respond by pressing a key corresponding to the orientation of the dots. There were three possible emotional pairings of the faces shown – 1. Neutral-neutral 2. Happy-neutral 3. Fearful-neutral. If participants had an attentional bias towards negative stimuli, they would respond faster when the dots appear in the location previously occupied by a fearful face than when the dots appeared in the other location. The emotional faces (happy and fearful) appeared in both locations at equal frequency. The faces shown were photographs of 20 individuals with different facial expressions taken from the JACFEE/JACNeuF sets of facial expressions [4]. Data was analysed by calculating attentional vigilance scores for each participant from mean reaction times. Namely, comparing between congruent trials, where probes appears in the same location as the emotional face, and incongruent trials, where probes appeared in the opposite position. After reaction times for incorrect trials were removed, and reaction time trials trimmed between 0.2-1.2ms, attentional vigilance scores were calculated for each participant by subtracting the mean reaction time from trials when probes appeared in the same position as the emotional face (congruent trials) from trials when probes appeared in the opposite position to the emotional face (incongruent trials). Positive values reflect attention towards the emotional face (vigilance) and negative values reflect attention away from the emotional face (avoidance).

In the Emotional Go/No-Go task, participants respond (go) or withhold responses (no-go) according to rules which change over time (e.g., instructions: do not press the button if you see a blue/yellow image) while being exposed to emotional distractors (fearful or happy faces, or control images) (as per https://www.nature.com/articles/s41467-024-50394-x). This task measured behavioural inhibition under affective interference (positive [happy faces], aversive/negative [fearful faces], and neutral distractors) where accuracy of inhibited response to no-go trials (response inhibition), accuracy, and response time for go trials (an index of impulsivity) were the non-model outcome measures. The block design of the task allowed for analysis of set-shifting effects (executive shifting for task condition rule changes) on accuracy and response time.

The DDM was derived from past work by Colwell where the model was validated (including parameter recovery and posterior predictive checks). The DDM contain parameters which describe distinct aspects of evidence accumulation throughout the task: 1) drift rate (split by stimulus: go or no-go trials) which refers to the efficiency of convergence toward the choice boundaries; 2) boundary separation which describes the magnitude of evidence required before reaching a decision; 2) non-decision time which describes the sensory and motor encoding/planning period prior to evidence accumulation; 4) initial choice bias which describes the initial bias toward the upper or lower choice boundary at the start of evidence accumulation; 5) drift criterion describes inherent biases within the direction or rate of evidence accumulation. The model uses gsquare optimisation to estimate the best fitting model for each participant within the following quantile bins: : 0.1, 0.3, 0.5, 0.7 and 0.9.

We also planned to undertake the Probabilistic Instrumental Learning Task (PILT) to assess reward learning and assessment. Unfortunately, there was an error in task delivery for a proportion of participants, and thus the data from this task was not suitable for analysis.

##### Outliers:

As per standard operating procedure, for the ETB tasks (FERT, ECAT, FDOT, EMEM), trials were excluded if reaction times were less than 200 ms, and greater than laboratory protocols for individual tasks in potentially depressed populations (10000 ms for the FERT; 3000ms for the ECAT and EMEM due to task design). FDOT trials were trimmed between 200ms and 1200ms. Across all tasks, if a participant’s performance met the criteria as an extreme outlier (data lying at more than three times the participants’ interquartile range above their third or below their first quartile, or less than 50% accuracy for the FDOT), or there were other concerns, the analysis was repeated with and without their data included as a sensitivity analysis. Any participants excluded from analysis are identified in the results for each task.

#### Participant Payment:

Participants were reimbursed £125 for their time upon completion of their participation in the research. If they did not complete the study, they will be given a pro-rata amount to recompense the time spent. Reasonable travel expenses for any visits were reimbursed on production of receipts, or a mileage allowance provided as appropriate.

###### ***Table S1: Exclusion criteria for the Progress Study***

Any current Axis 1 DSM-5 psychiatric disorder

Any previous episode of a severe mental illness, other than Depressive Disorder. Comorbid Anxiety disorders will be allowed, but not OCD or PTSD.

A first degree relative diagnosed with Bipolar Affective Disorder Type 1 or Schizophrenia

Body Mass Index outside the range of 18 to 33 inclusive

Any significant current medical condition likely to interfere with conduct of the study or analysis of data

Current use of psychoactive and / or medically significant medication as judged by a study medic, whether prescribed or bought over the counter (the contraceptive pill, the Depo-Provera injection or the progesterone implant will not result in exclusion)

Ongoing psychopharmacological treatment for depression, including hypnotics (psychotherapy will be allowed as long as not newly-started in the last 6 weeks)

High consumption of licit substances to an extent that would make complying with study protocol challenging (including alcohol, caffeine, nicotine)

Past history of dependence to illicit substances, and any consumption of illicit substances in the three months prior to the study

Currently pregnant or breast feeding

Current, or a significant history of, gastro-intestinal disorder or irritable bowel syndrome

Known lactase deficiency or any other problem absorbing lactose, galactose, or glucose

Participation in a study that involves the use of a medication or novel vaccine within the last three months

Participation in a study using the same tasks in the last two years

Any physical (including visual and auditory) or language impairment that would make complying with the study protocol challenging

#### Statistical Plan

##### Data Cleaning

- Will be performed prior to unblinding
- Outliers will be excluded on a per task basis
- For all task data, cut-off thresholds will be determined based on visually inspecting plots, examining thresholds for:
  - Trials with unusually low or high response times
  - Proportion of missing trials per participant
  - Unusually low mean accuracy (or equivalent outcome) per participant
  - Unusually low or high mean reaction time per participant
- For all self-report/questionnaire data, extreme outliers indicating invalid data entry will be determined based on visually inspection.

Data quality meeting occurred on 29^th^ November 2023. Final agreement on outliers 21^st^ December 2023.

SAP and data quality decisions occurred with AG, AdeC, CH whilst blind to allocation. SAP agreed and uploaded to OSF prior to code break.

##### Task Analysis

| Task | Outcomes | Analysis |
| --- | --- | --- |
| Auditory Verbal Learning Task (AVLT)  Recalling words read aloud | **Number of words recalled** – List A immediate recall trials | Repeated measures analyses of covariance (ANCOVAs):   - Between-subject factor – 2 levels: Treatment group (prucalopride or placebo) - Within-subject factor – 5 levels: Trial (List A immediate recall trials 1-5) - Covariate: Timepoint (Baseline vs Research visit) |
|  | **Number of words recalled** – List A short delay  **Number of words recalled** – List A long delay | Repeated measures analyses of covariance (ANCOVAs):   - Between-subject factor – 2 levels: Treatment group (prucalopride or placebo) - Within-subject factor – 2 levels: List (List A short and long delay trials) - Covariate: Timepoint (Baseline vs Research visit) |
|  | **Number of intrusions (words incorrectly recalled)** across List A acquisition trials | Repeated measures analyses of covariance (ANCOVAs):   - Between-subject factor – 2 levels: Treatment group (prucalopride or placebo) - Covariate: Timepoint (Baseline vs Research visit) |
|  | **Number of repetitions (words repeated within same trial)** across List A acquisition trials |  |
|  | **Number of words recalled** – List B recall |  |
|  | **Number of hits and false alarms in the delayed recognition test** |  |
| **N-back Task**  Recognition of ‘target’ letters | **% Accuracy of target words recalled** | Repeated measures analyses of covariance (ANCOVAs):   - Factors – 2 levels: Treatment group (prucalopride or placebo) - Covariate: Timepoint (Baseline vs Research visit) |
|  | **% Accuracy of non-target words recalled** |  |
|  | **Reaction time (ms)** for trials with correct responses |  |
| **Emotional Go/No-Go Task** | **% Accuracy of go targets** | Repeated measures analyses of covariance (ANCOVAs):   - Between-subject factor – 2 levels: Treatment group (prucalopride or placebo) - Covariate: Timepoint (Baseline or Research visit) |
|  | **% Accuracy of no-go targets** |  |
|  | **Reaction time (ms)** for trials with correct responses |  |
| **Digit Symbol Substitution Test (DSST)** | **Number of correct symbols** | Repeated measures analyses of variance (ANCOVAs):   - Between-subject factor – 2 levels: Treatment group (prucalopride or placebo) - Covariate: Timepoint (Baseline or Research visit) |
| **Trail-making task** | **Time to completion (TMT-A)** | Independent samples t- tests comparing groups (data obtained at Research visit only) |
|  | **Time to completion (TMT- B)** |  |
|  | **Ratio between performance in part B:part A** |  |
| **Facial Expression Recognition Task (FERT)**  Recognition of computer-based positive and negative facial expressions | **Unbiased hit rate:** a measure of emotion identification accuracy which accounts for response bias i.e. any general tendency to identify the emotion when it is not present. Calculated as proportion of correct hits* (number of hits/all hits and misses) for each facial expression category. % correct and response bias will also be reported individually | Repeated measures analyses of variance (ANOVAs) (only conducted at Research visit):   - Between-subject factor – 2 levels: Treatment group (prucalopride or placebo) - Within-subject factor – 7 levels: (Fear, anger, happy, surprise, disgust, sad, neutral) |
|  | **Misclassifications:** Number of responses to each facial expression category classified as another category i.e. identifying a fearful face as surprised |  |
|  | **Reaction time (ms)** for trials with correct responses |  |
| **Emotional Categorisation Task (ECAT)**  Categorisation of emotional words | **% Accuracy –** words correctly identified as positive or negative | Mixed model analyses of variance (ANOVAs) (only conducted at Research visit):   - Between-subject factor – 2 levels: Treatment group (prucalopride or placebo) - Within-subject factor – 2 levels: Word valence (positive or negative) |
|  | **Reaction Time** |  |
| **Emotional Recall Task (EREC)**  Recall of emotional words from ECAT | **Number of hits** (words recalled correctly) |  |
|  | **Number of false alarms** (words recalled incorrectly) |  |
| **Emotional Recognition Task (EMEM)** | **Number of hits** (words recognised correctly) |  |
|  | **Number of false alarms** (words recognised incorrectly) |  |
|  | **Reaction time** |  |
| **Faces Dot Probe Task (FDOT)** | **Number of hits** (recognised in correct orientation) | Mixed model analyses of variance (ANOVAs) (only conducted at Research visit):   - Between-subject factor – 2 levels: Treatment group (prucalopride or placebo) - Within-subject factor – 2 levels: Symbol orientation (horizontal or vertical) - Within-subject factor – 2 levels: Vigilance (masked or unmasked) |
|  | **Number of false alarms** (recognised incorrectly) |  |
|  | **Reaction time** |  |

### Supplementary Results

#### **Questionnaire results**

###### ***Figure S1: CONSORT 2025 flow diagram***

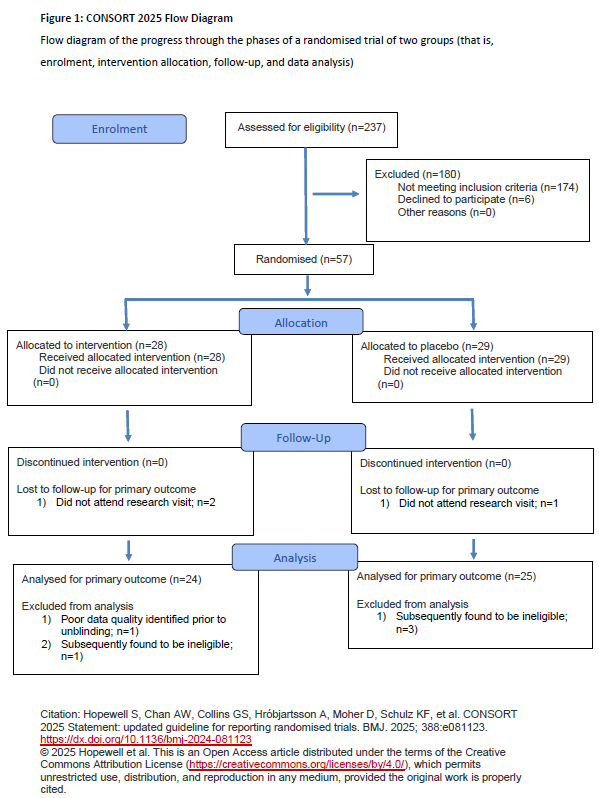

###### ***Table S2: Side effects questionnaire mean scores (% of participants in interventional group reporting presence / absence in prior week)***

|  | ***PRE-INTERVENTION (%)*** | | ***POST-INTERVENTION (%)*** | | ***Time * Group interaction on ANOVA*** |
| --- | --- | --- | --- | --- | --- |
|  | ***Placebo*** | ***Prucalopride*** | ***Placebo*** | ***Prucalopride*** |  |
| **Headache** | 12 | 4 | 12 | 12 | F(1,47)=1.04, p=0.31 |
| **Abdominal Pain** | 4 | 8 | 12 | 8 | F(1,47)=0.96, p=0.33 |
| **Nausea** | 4 | 4 | 16 | 4 | F(1,47)=1.79, p=0.19 |
| **Diarrhoea** | 0 | 0 | 16 | 12 | F(1,47)=0.12, p=0.733 |
| **Decreased Appetite** | 12 | 12 | 4 | 24 | **F(1,47)=5.42, p=0.024** |
| **Dizziness** | 0 | 0 | 8 | 0 | F(1,47)=2.00, p=0.16 |
| **Vomiting** | 0 | 0 | 0 | 0 | NA (zero in all pts) |
| **Flatulence** | 8 | 4 | 20 | 12 | F(1,47)=0.12, p=0.731 |
| **Fatigue** | 48 | 20 | 44 | 12 | F(1,47)=0.07, p=0.79 |
| **Gastrointestinal sounds** | 12 | 8 | 12 | 24 | F(1,47)=2.18, p=0.15 |

###### Supplementary results for side effects

Reports of diarrhoea and flatulence increased or were borderline increased for both groups from baseline [main effect of time for diarrhoea: F(1,47)=7.81, p=0.007, np^2^=0.14 ; and flatulence: F(1,47)=3.68, p=0.06, np^2^=0.07], but the group*symptom interaction was not significant [group*time interaction diarrhoea: F(1,47)=0.12, p=0.733, np^2^<0.01; and flatulence: F(1,47)=0.12, p=0.731, np^2^<0.01]. Fatigue was higher in the placebo group at both time points [main effect of group: F(1,47)=8.55; p=0.005; group*time interaction: F(1,47)=0.07, p=0.79].

###### Figure S2: QIDS scores at post-intervention versus PHQ9 baseline scores converted to QIDS equivalent across groups

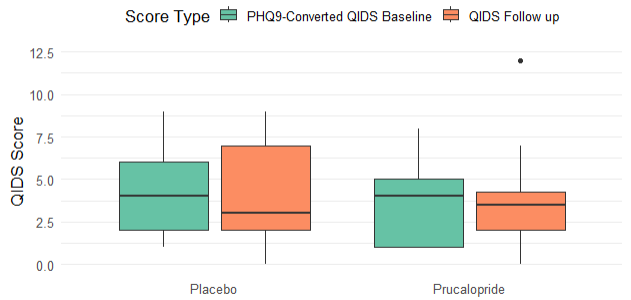

#### AVLT

N = Placebo 25, Prucalopride 24

We adjusted data relating to a very likely error in recording the scores for one participant in the placebo group in the AVLT post-intervention: who scored 10 on trial 3 and 5 but was recorded as scoring 0 on trial 4 in List A. We assumed that the recording of 0 was an error for trial 4, and therefore imputed 10 as the score for trial 4 in main analyses. We also ran a sensitivity analysis using the original data, as reported in the main paper (see below).

###### Table S3: AVLT mean scores (SD) for the Free recall list A (max score 15)

| **Timepoint** | **Intervention** | **Block (Free recall list A)** | **Mean score** | **SD** |
| --- | --- | --- | --- | --- |
| **Baseline** | **Placebo** | 1 | 8.16 | 2.25 |
|  |  | 2 | 11.4 | 2.45 |
|  |  | 3 | 12.6 | 2.31 |
|  |  | 4 | 13.0 | 2.00 |
|  |  | 5 | 12.9 | 2.22 |

|  | **Prucalopride** | 1 | 6.75 | 1.67 |
| --- | --- | --- | --- | --- |
|  |  | 2 | 10.46 | 1.67 |
|  |  | 3 | 11.9 | 1.82 |
|  |  | 4 | 12.9 | 1.21 |
|  |  | 5 | 13.1 | 1.70 |
| **Post-intervention** | **Placebo** | 1 | 7.96 | 2.44 |
|  |  | 2 | 11.1 | 2.87 |
|  |  | 3 | 12.4 | 1.94 |
|  |  | 4 | 12.8 | 2.09 |
|  |  | 5 | 13.0 | 1.83 |
|  | **Prucalopride** | 1 | 8.75 | 1.78 |
|  |  | 2 | 11.3 | 1.86 |
|  |  | 3 | 12.7 | 1.63 |
|  |  | 4 | 13.4 | 1.44 |
|  |  | 5 | 13.8 | 1.44 |

###### Supplementary results for the AVLT

*Sensitivity analyses for main effect:*

i. When we did not account for baseline scores [F(1,220)=4.97, p=0.027]
ii. When we used raw data without adjustment for an error in scoring for one placebo participant [F(1,226)=9.08, p=0.003] (see above)
iii. When we included native language as a covariate [F(1,224)=8.16, p=0.005]
iv. When we included baseline mood score (PHQ-9) and subjective cognition (PDQ-20) as a covariate [F(1,222)=8.31, p=0.004]

*Analyses checking significance of version order*

ANOVA demonstrating that version order had a statistically significant effect on mean List A score (across the first 5 trials): F(1,266) = 4.75, p=0.034

*Other analyses*

ANOVA for other parts of the AVLT showing no group-level significance on the following scores:

Intrusions: [F(1,42)=0.96, p=0.33, np2 <0.01].

Repetitions: [F(1,42)=0.88, p=0.35, np2 <0.01].

List B: [F(1,42)=0.33, p=0.57, np2 <0.01].

###### ***Figure S3A and B: Mean numbers of words recalled across the placebo and prucalopride groups for all parts of the task, comparing baseline and post-intervention results***

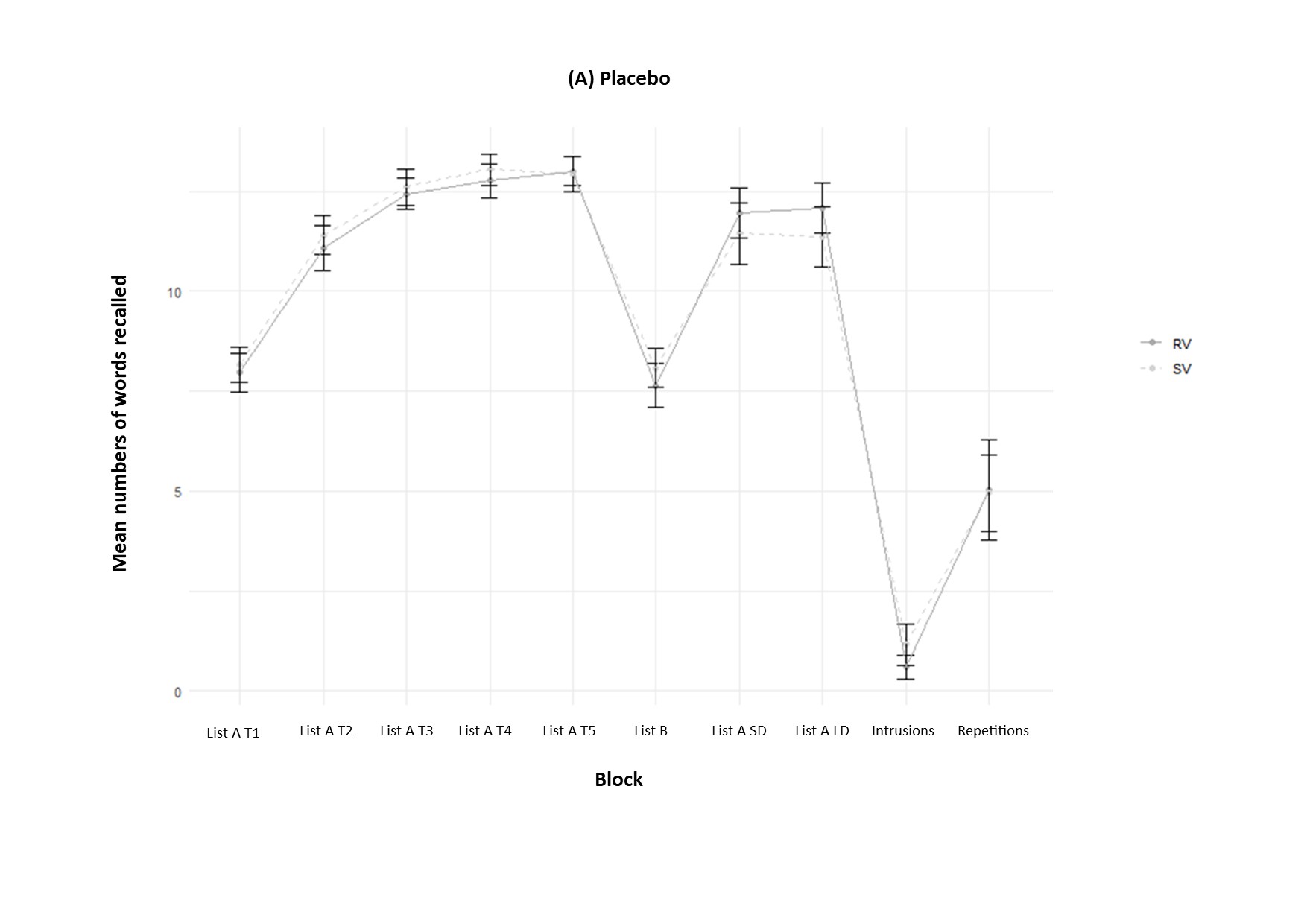

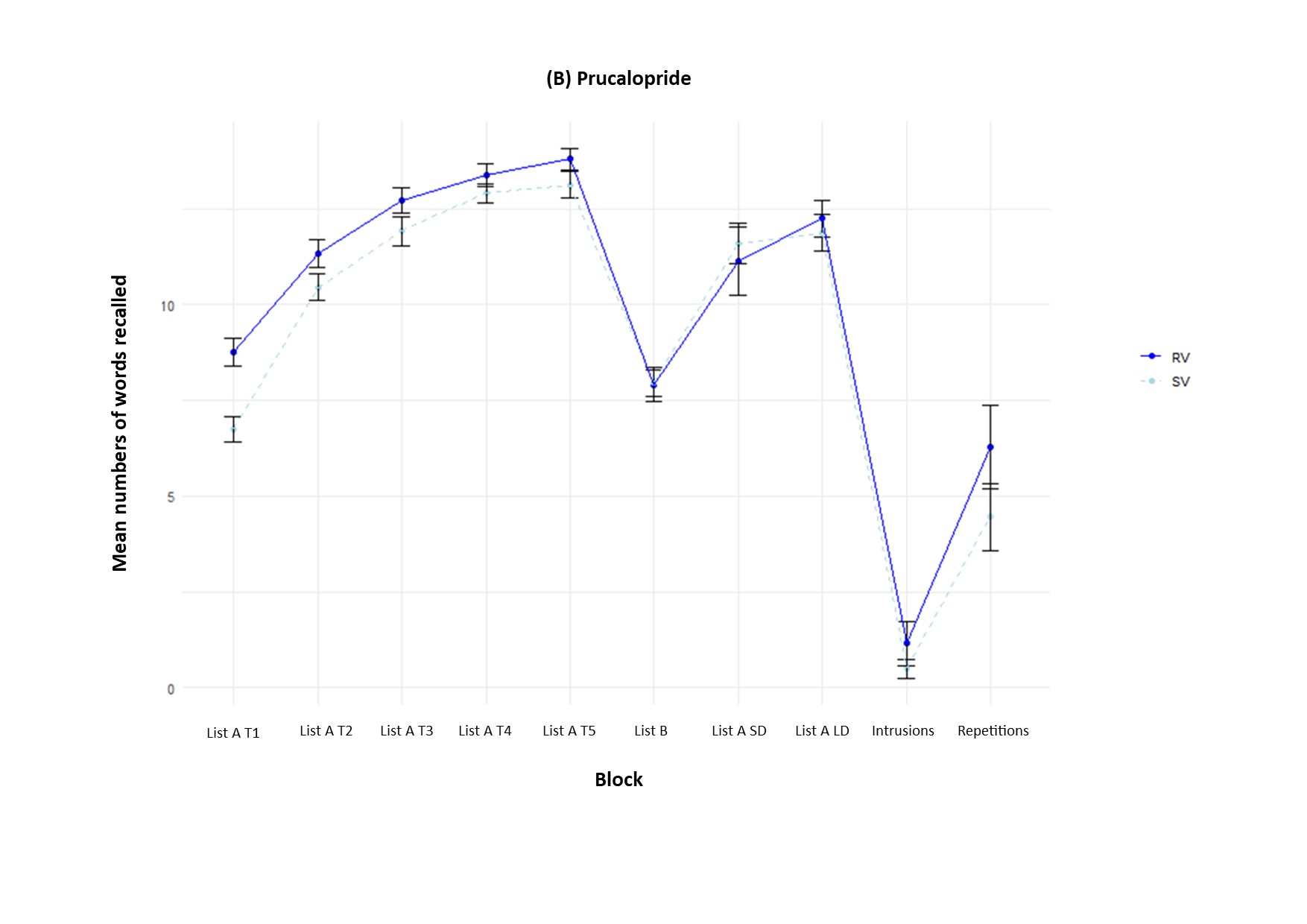

RV = research visit (post-intervention); SV = screening visit. Error bars show standard error of the mean.

###### ***Figure S3C: Mean numbers of words recalled across the placebo and prucalopride groups for intrusions and repetitions, comparing baseline and post-intervention results***

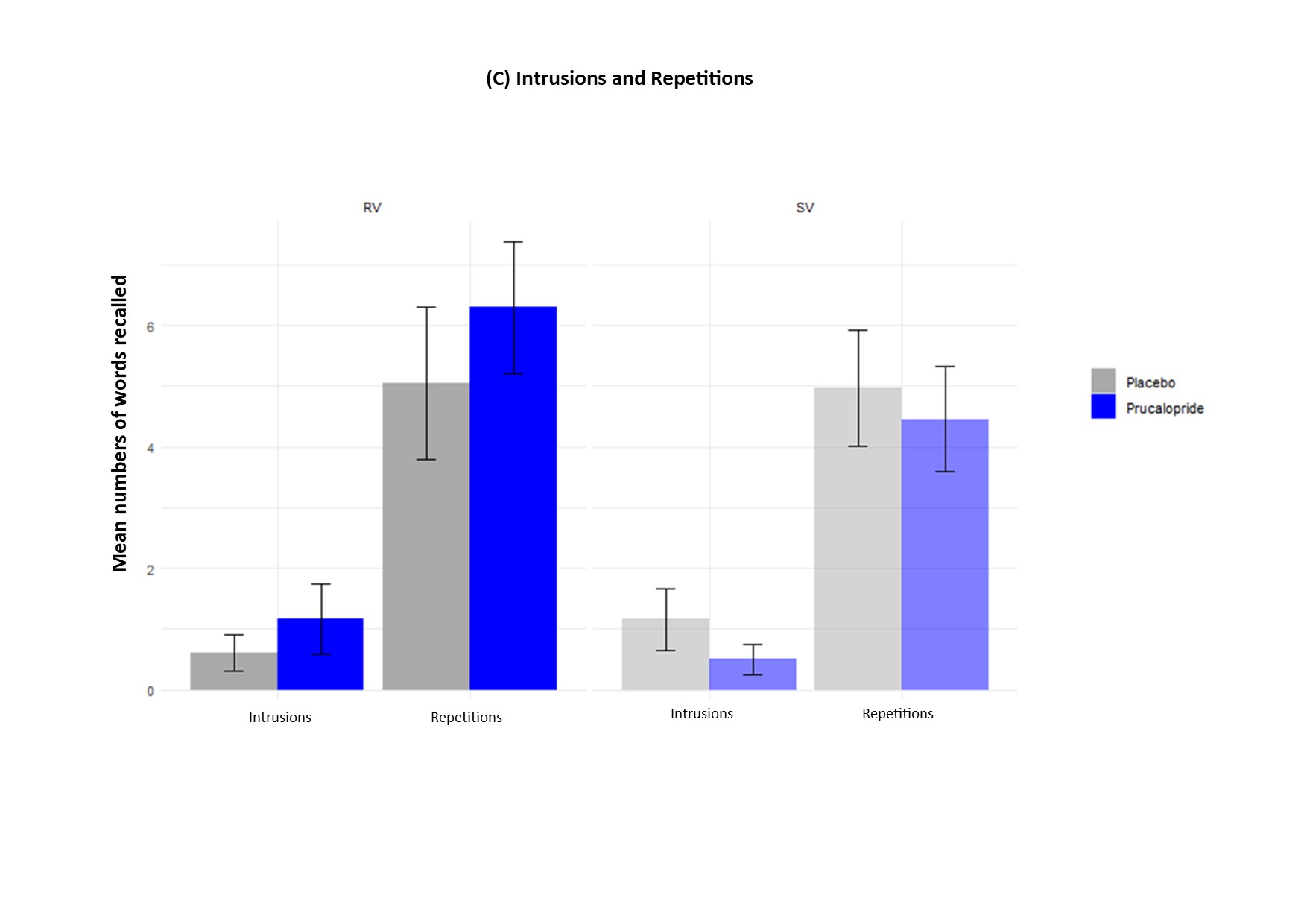

RV = research visit (post-intervention); SV = screening visit. Error bars show standard error of the mean.

#### N-back

N = Placebo 25, Prucalopride 23

###### Table S4: Mean accuracy (score) for the N-back

| **Group** | **n-back** | **Mean score** | **SD** |
| --- | --- | --- | --- |
| ***Placebo*** | 0 | 8.88 | 0.88 |
| ***Prucalopride*** | 0 | 9.04 | 0.48 |
| ***Placebo*** | 1 | 8.96 | 2.01 |
| ***Prucalopride*** | 1 | 9.48 | 1.08 |
| ***Placebo*** | 2 | 7.68 | 1.38 |
| ***Prucalopride*** | 2 | 8.17 | 0.78 |
| ***Placebo*** | 3 | 8.24 | 1.62 |
| ***Prucalopride*** | 3 | 8.09 | 1.88 |

###### Table S5: Mean reaction time (ms) for the N-back

| **Group** | **n-back** | **Mean score** | **SD** |
| --- | --- | --- | --- |
| ***Placebo*** | 0 | 573.70 | 253.43 |
| ***Prucalopride*** | 0 | 499.16 | 114.00 |
| ***Placebo*** | 1 | 678.20 | 326.38 |
| ***Prucalopride*** | 1 | 564.45 | 142.22 |
| ***Placebo*** | 2 | 752.80 | 274.57 |
| ***Prucalopride*** | 2 | 671.05 | 213.74 |
| ***Placebo*** | 3 | 815.74 | 244.74 |
| ***Prucalopride*** | 3 | 748.93 | 198.08 |

###### Supplementary results for the N-back

*Reaction time*

Main analyses for reaction time remained significant when baseline data was not included as a covariate [F(1,180)=4.45, p=0.036], and when the model included subjective cognition and mood scores at baseline [F(1,155)=9.32, p=0.003].

Reaction time analyses were borderline when we excluded a placebo participant who was an outlier for baseline and follow-up reaction time (reduced speed), but not for accuracy [F(1,175)=3.71, p=0.05].

*Accuracy*

Main analyses for accuracy were also unchanged when accounting for mood and subjective cognition scores at baseline [F(1,155)=2.10, p=0.15)].

Accuracy analyses were similar when a placebo participant who was an outlier for baseline and follow-up reaction time (reduced speed) but not for accuracy was excluded [F(1,175)=3.01, p=0.08)]

*Signal detection analyses*

ANCOVA for d’ and group including RT as covariate: F(1,44)=2.69, p=0.10, np2=0.05

###### ***Figure S4: d’prime versus reaction time for target hits comparing placebo and prucalopride groups***

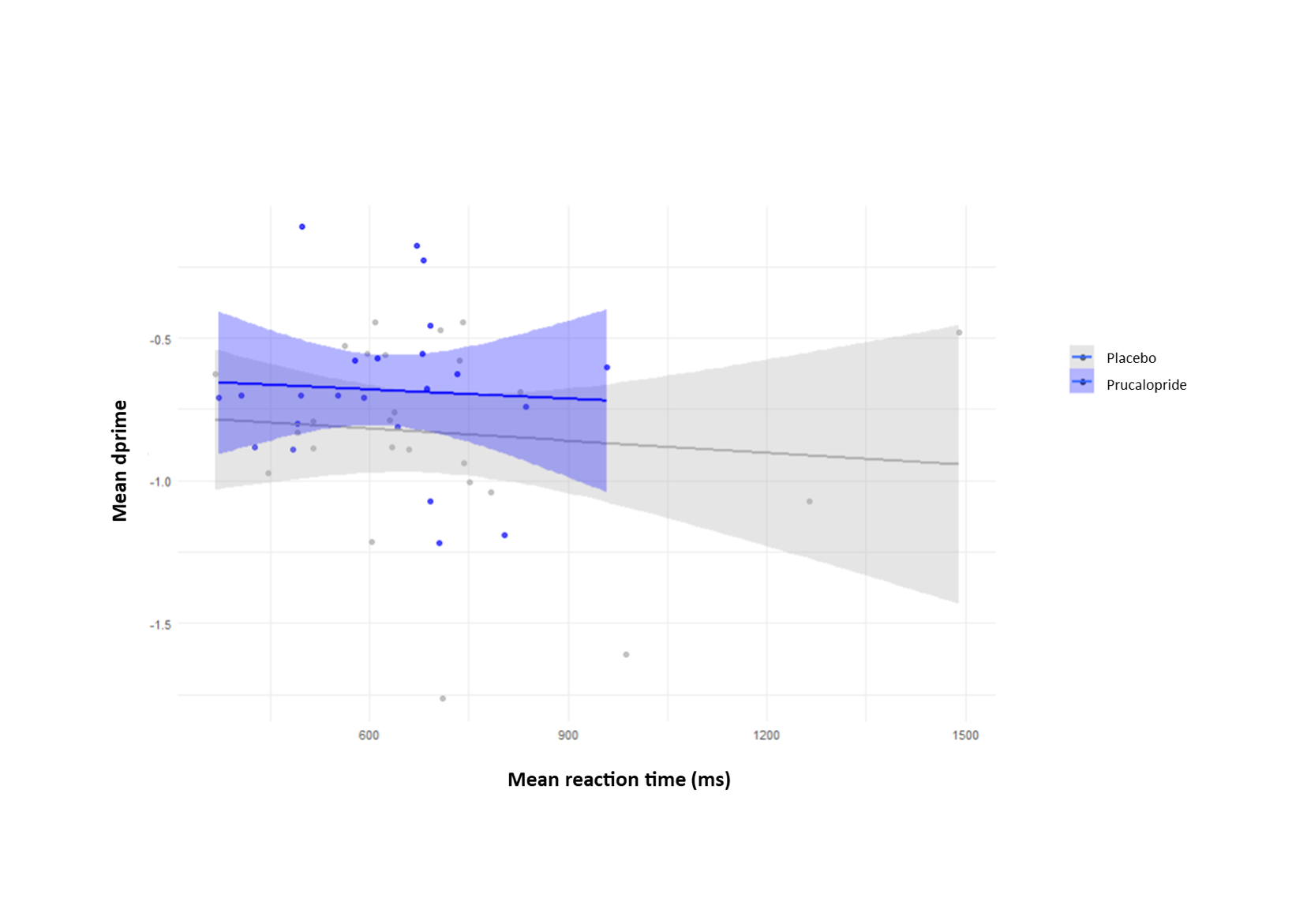

#### DSST

N = Placebo 25, Prucalopride 24

###### Table S6: Mean reaction time for the DSST

| **Group** | **Reaction time baseline (s; SD)** | **Reaction time post-intervention (s; SD)** |
| --- | --- | --- |
| ***Placebo*** | 50.2 (12.7) | 57.3 (12.7) |
| ***Prucalopride*** | 54.0s (13.5 | 57.6 (14.0) |

###### Supplementary results for the DSST

We assessed the impact of prucalopride allocation on time to complete the DSST. There was no group level significance for main analyses (adjusted for baseline DSST time and version order), or in sensitivity analyses as follows:

*Adjusted for baseline DSST and version order:*

DSST Group difference: F(3,45)=-0.88, p=0.38; placebo baseline mean (SD) 50.2 (12.7), placebo post-intervention mean (SD) 57.3 (12.7); prucalopride baseline mean (SD) 54.0 (13.5), prucalopride post-intervention mean (SD) 57.6 (14.0)

*When not adjusting for baseline DSST:*

DSST Group difference: F(1,46)=0.009, p=0.92

*Also adjusted for age and years of education:*

DSST Group difference: F(5,43)=-0.92, p=0.36

*Also adjusted for baseline PDQ20 and PHQ9:*

TMTB-TMTA Group difference: F(5,43)=-1.01, p=0.32

###### ***Figure S5: Time to complete the DSST at baseline and research (post-intervention) visits comparing prucalopride and placebo allocation and version administered***

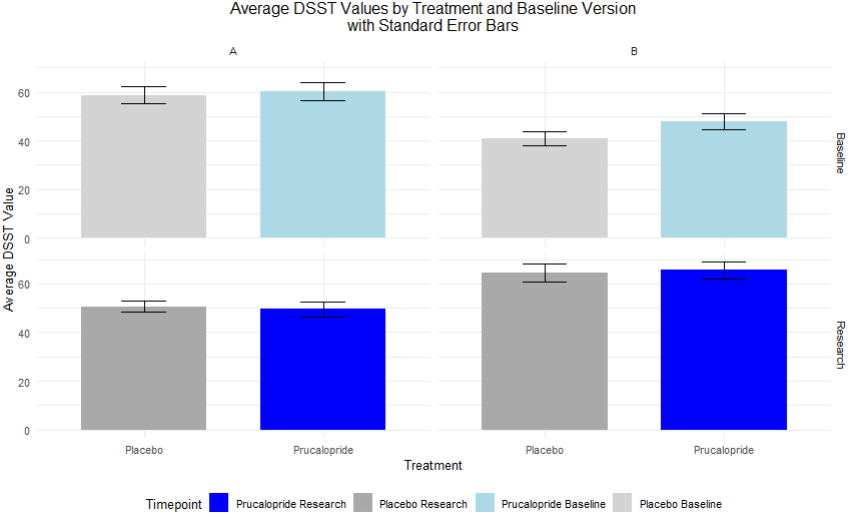

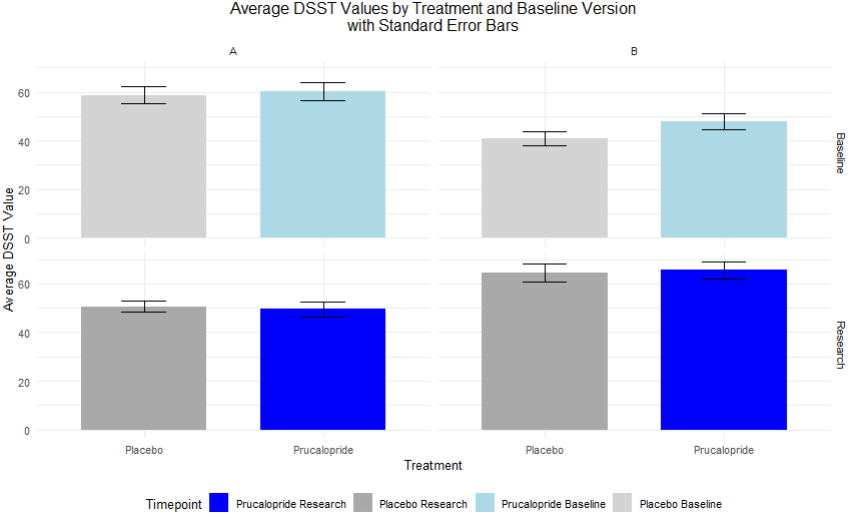

#### TMT

N = Placebo 24, Prucalopride 22

We assessed the significance of prucalopride allocation compared to placebo on time to complete the TMT (parts A and B). Due to missing data, 24 placebo participants had TMT data available for analysis, and 22 prucalopride participants.

###### Table S7: Mean reaction time for the TMT

| **Group** | **TMTB-TMTA (s; SD)** | **TMTB (s; SD)** | **TMTA (s; SD)** |
| --- | --- | --- | --- |
| ***Placebo*** | 30.8 (21.9), | 57.8 (22.3) | 26.9 (10.2), |
| ***Prucalopride*** | 27.8 (12.5) | 54.7 (14.5) | 26.9 (9.2) |

###### Supplementary results for the TMT

TMTA Group difference: t(44.0)=0.007, p=0.99; placebo mean (SD) 26.9s (10.2), prucalopride mean (SD) 26.9s (9.2)

- *Measures processing and motor speed and visual scanning*

TMTB Group difference: t(39.9)=0.56, p=0.58; placebo mean (SD) 57.8s (22.3), prucalopride mean (SD) 54.7s (14.5)

- *Measures cognitive flexibility and executive functioning (but without accounting for differences in processing speed)*

TMTB minus TMTA Group difference: t(37.1)=0.58, p=0.56; placebo mean (SD) 30.8s (21.9), prucalopride mean (SD) 27.8s (12.5)

- *Purer measure of executive functioning accounting for differences in processing speed*

*Above adjusted for age and years of education:*

TMTB-TMTA Group difference: F(3,42)=-0.34, p=0.74

*Above adjusted for baseline PDQ20 and PHQ9:*

TMTB-TMTA Group difference: F(3,42)=-0.46, p=0.65

###### ***Figure S6: Time to complete the TMTB (after subtracting TMTA) comparing prucalopride and placebo allocation***

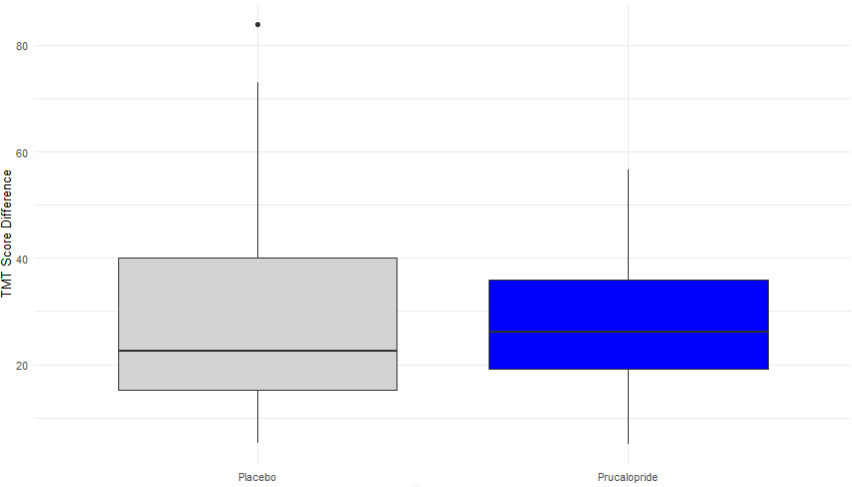

Box and whisker plot with whiskers demonstrating 1.5x upper and lower interquartile range

#### FERT

N = Placebo 25, Prucalopride 23

1 Prucalopride participant did not have data collected as they had recently undertaken the Emotional Test Battery.

###### Table S8: Percentage mean accuracy (SD) and mean reaction time (SD) for the FERT

|  | **Emotion** | **Mean accuracy (%)** | **SD** | **Mean reaction time (ms)** | **SD** |
| --- | --- | --- | --- | --- | --- |
| **Prucalopride** | ***ANGER*** | 57.40000 | 16.273701 | 1847.917 | 519.7726 |
|  | ***DISGUST*** | 47.10000 | 14.043830 | 1821.864 | 394.1998 |
|  | ***FEAR*** | 30.90000 | 14.805686 | 2277.883 | 1276.4210 |
|  | ***HAPPY*** | 71.50000 | 10.968705 | 1450.551 | 224.8050 |
|  | ***SAD*** | 58.00000 | 13.268069 | 1595.970 | 321.8322 |
|  | ***SURPRISE*** | 73.70000 | 7.221207 | 1515.221 | 269.9253 |
| **Placebo** | ***ANGER*** | 53.80435 | 18.182621 | 1546.992 | 302.2142 |
|  | ***DISGUST*** | 41.19565 | 12.405771 | 1581.534 | 355.2165 |
|  | ***FEAR*** | 26.52174 | 14.533252 | 1843.610 | 387.7698 |
|  | ***HAPPY*** | 71.95652 | 9.533333 | 1331.044 | 262.9369 |
|  | ***SAD*** | 53.04348 | 12.153295 | 1485.950 | 262.9424 |
|  | ***SURPRISE*** | 67.93478 | 9.730814 | 1373.900 | 358.9745 |

###### Supplementary results for the FERT

Results for main effect of group for primary analyses remained significant for accuracy [F(1,234)=9.29, p=0.0026] and reaction time [F(1,233)=13.3, p=0.0003] when scores for subjective cognition (PDQ20) and depressive symptom (PHQ-9) questionnaires at baseline were incorporated as covariates into the model.

When averaged across all emotions, prucalopride had a borderline main effect of group regarding misclassifications: [F(1,45)=3.87, p=0.05, np2=0.09 (0.00-1.00)].

#### FDOT

N = Placebo 24, Prucalopride 22

###### ***Table S9: Mean values for vigilance (SD) for the FDOT***

| **Group** | **Emotion** | **Mask** | **Mean vigilance** | **SD** |
| --- | --- | --- | --- | --- |
| **Placebo *FEAR*** | | Masked | -0.024372645 | 0.109 |
|  |  | Unmasked | 0.019585421 | 0.132 |
| **Placebo *HAPPY*** | | Masked | 0.008516653 | 0.111 |
|  |  | Unmasked | -0.017581067 | 0.108 |
| **Prucalopride *FEAR*** | | Masked | -0.010633539 | 0.134 |
|  |  | Unmasked | -0.005894571 | 0.133 |
| **Prucalopride *HAPPY*** | | Masked | -0.011922355 | 0.138 |
|  |  | Unmasked | -0.004708423 | 0.144 |

###### Supplementary results for the FDOT:

Attentional vigilance results for group*emotion*mask interaction were unchanged when PHQ9 and PDQ were added into the model [F(1,662)=4.23, p=0.04] and when two participants excluded for less than 50% accuracy were included [F(1,695)=4.65, p=0.03].

Results for reaction time were similarly unchanged when adding PHQ and PDQ added into model [F(1,727)=9.49, p=0.0021].

Including data from two participants excluded for less than 50% accuracy did render the result non-significant [F(1,765)=4.95, p=0.34], but these participants were excluded prior to unblinding and analyses being undertaken due to their poor accuracy across the task (less than chance) indicating lack of engagement.

Calculating proportional difference scores ((incongruent RT / congruent RT) / mean RT) instead of vigilance had little impact on results: Group*emotion*mask interaction [F(1,686)=4.02, p=0.045].

###### ***Figure S7: Results for the Facial Dot Probe Task (FDOT) across the prucalopride and placebo groups***

**(A)**

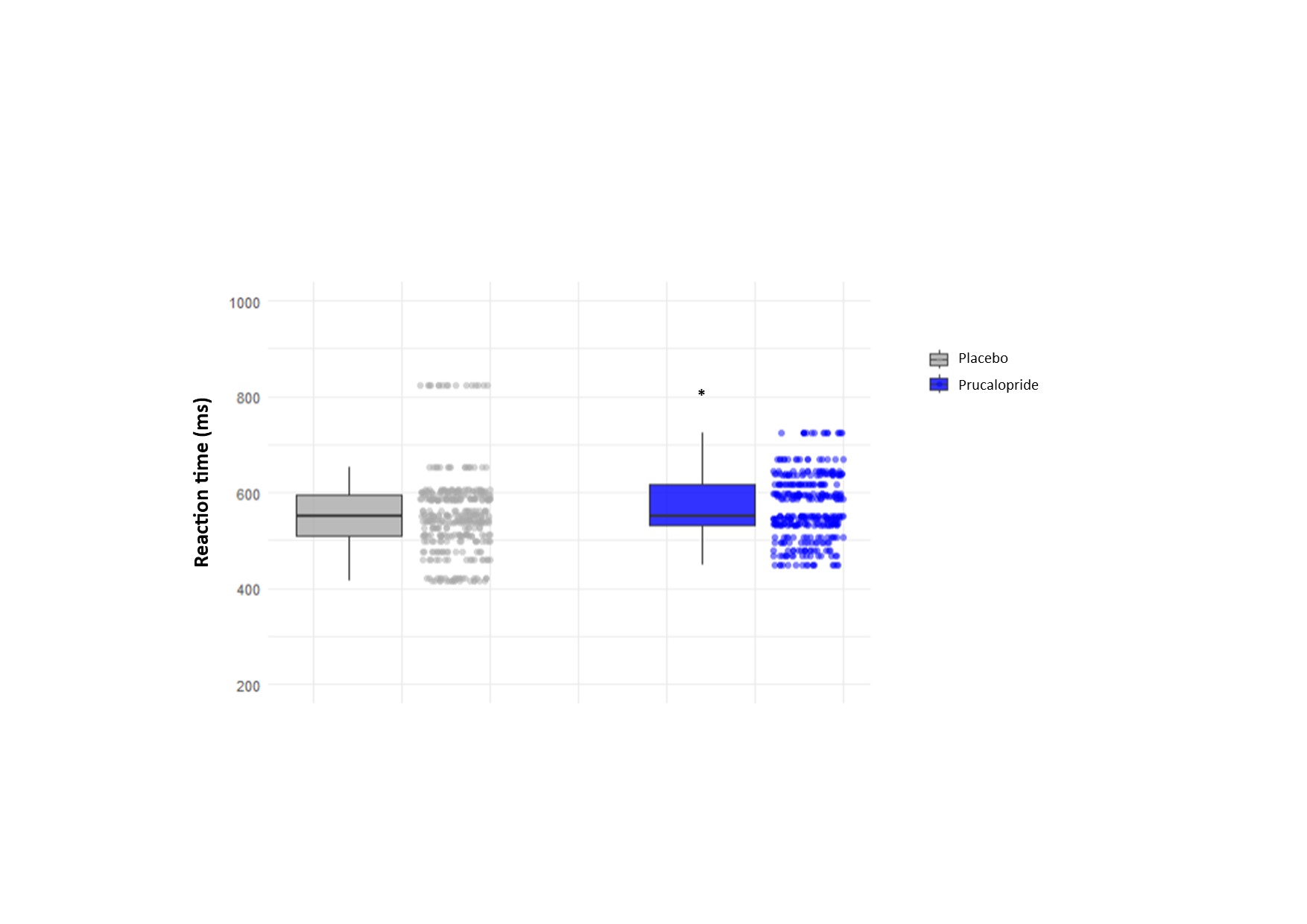

**(B)**

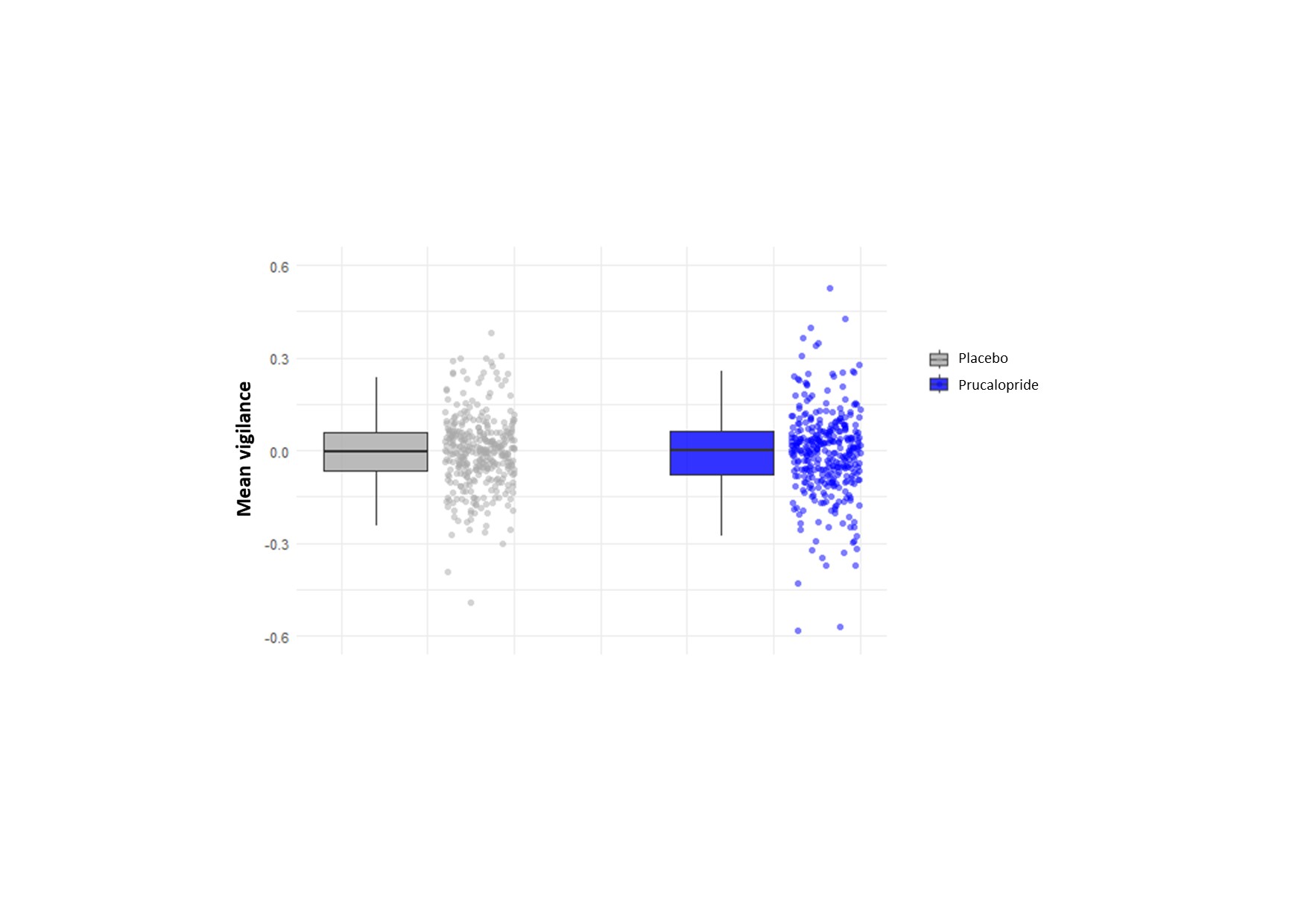

**(Ci)**

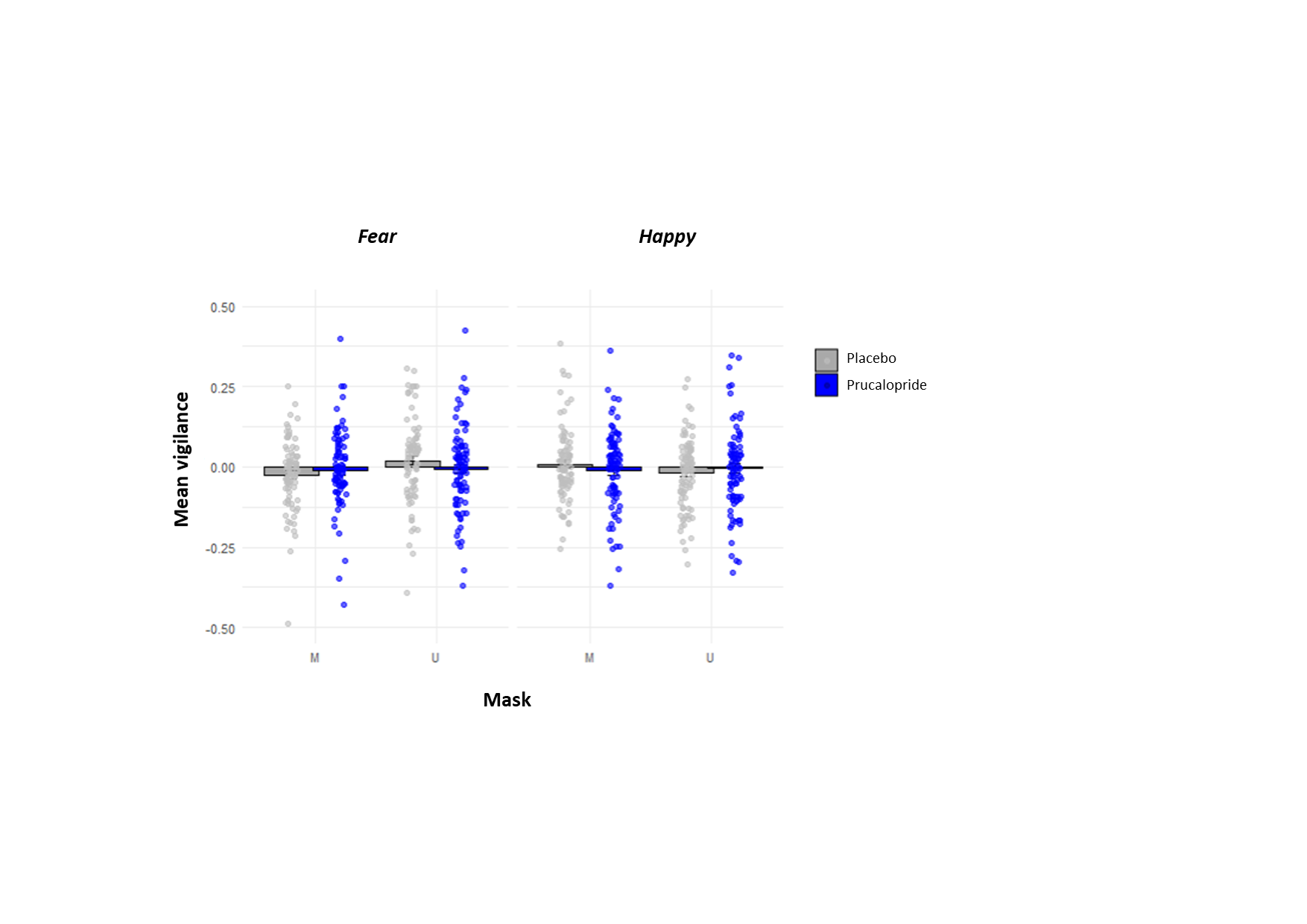

**(Cii)**

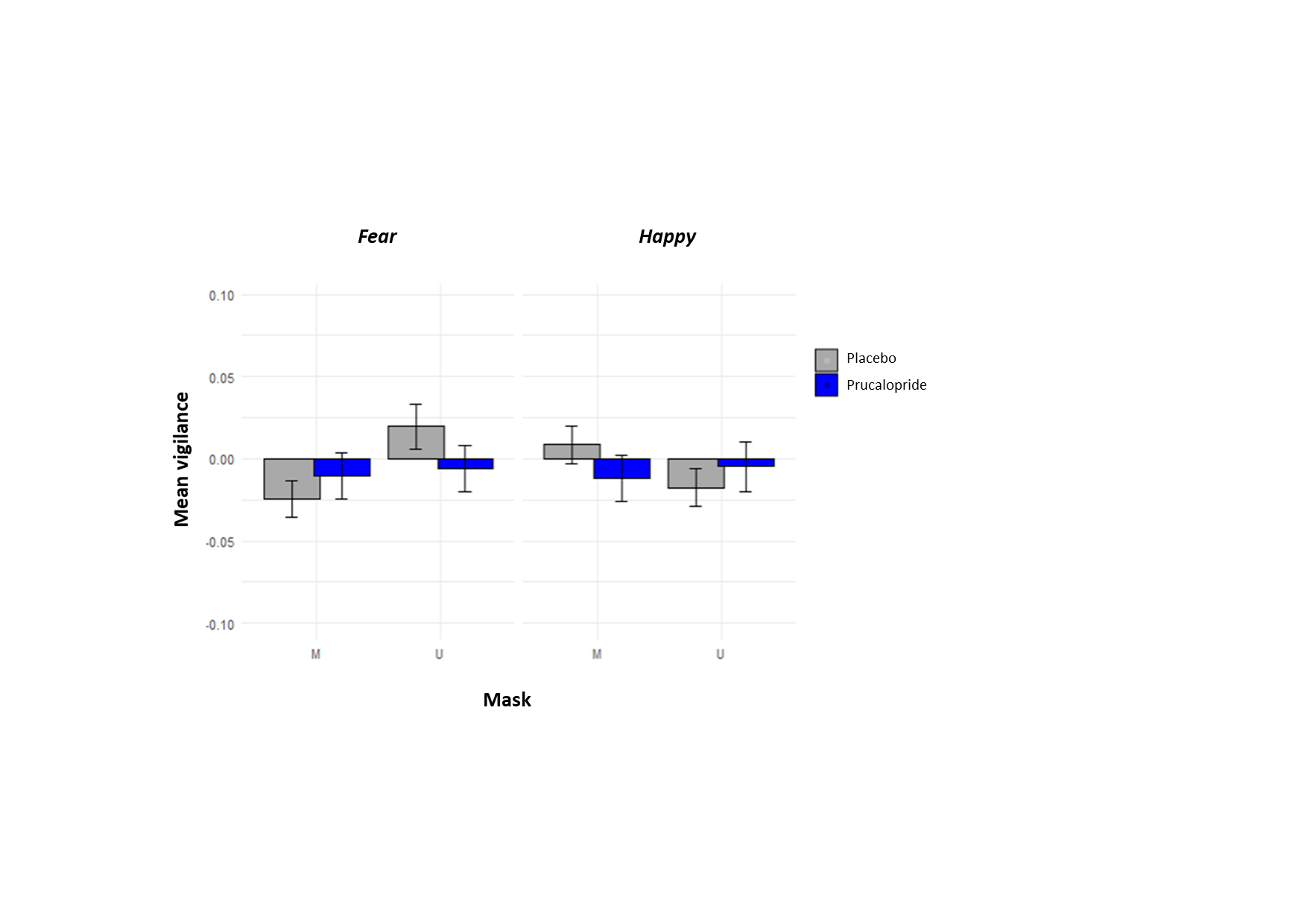

(A) Mean reaction time (ms) across the task compared by group; (B) Mean vigilance compared by group. Central line represents the median, the box represent the interquartile range, and bars the spread of the data. * represents statistical significance at p=0.05; (C) Mean vigilance across groups when stratified by mask / unmasked stimuli and fearful / happy stimuli (i) with data points (ii) without data points. Error bars represent standard error of the mean.

#### EGNG

N = Placebo 23, Prucalopride 22

###### ***Figure S8A and B: Results of the EGNG task in prucalopride and placebo groups: accuracy of no-go responses (response inhibition) in context of set shifting and accuracy go responses in context of affective interference***

**(A)	(B)**

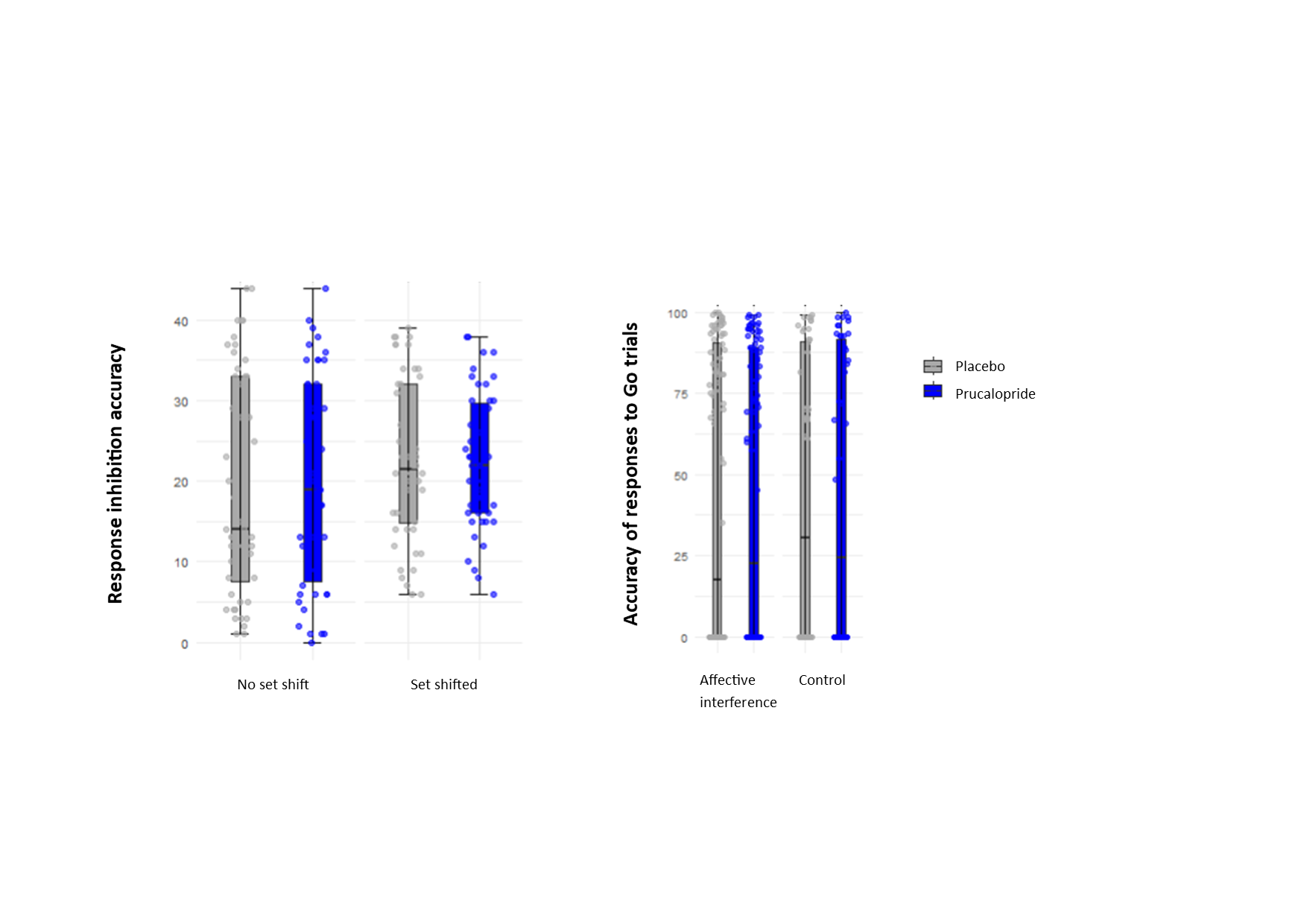

(A) Response inhibition (accuracy of no presses) by set shifting compared across groups; (B) Accuracy of Go responses by affective interference across groups; Whiskers demonstrate 1.5 x interquartile range, central line represents the median.

###### ***Figure S8C: Results of the EGNG task in prucalopride and placebo groups: accuracy of no-go responses (response inhibition) and time to choice (reaction time) by trial type***

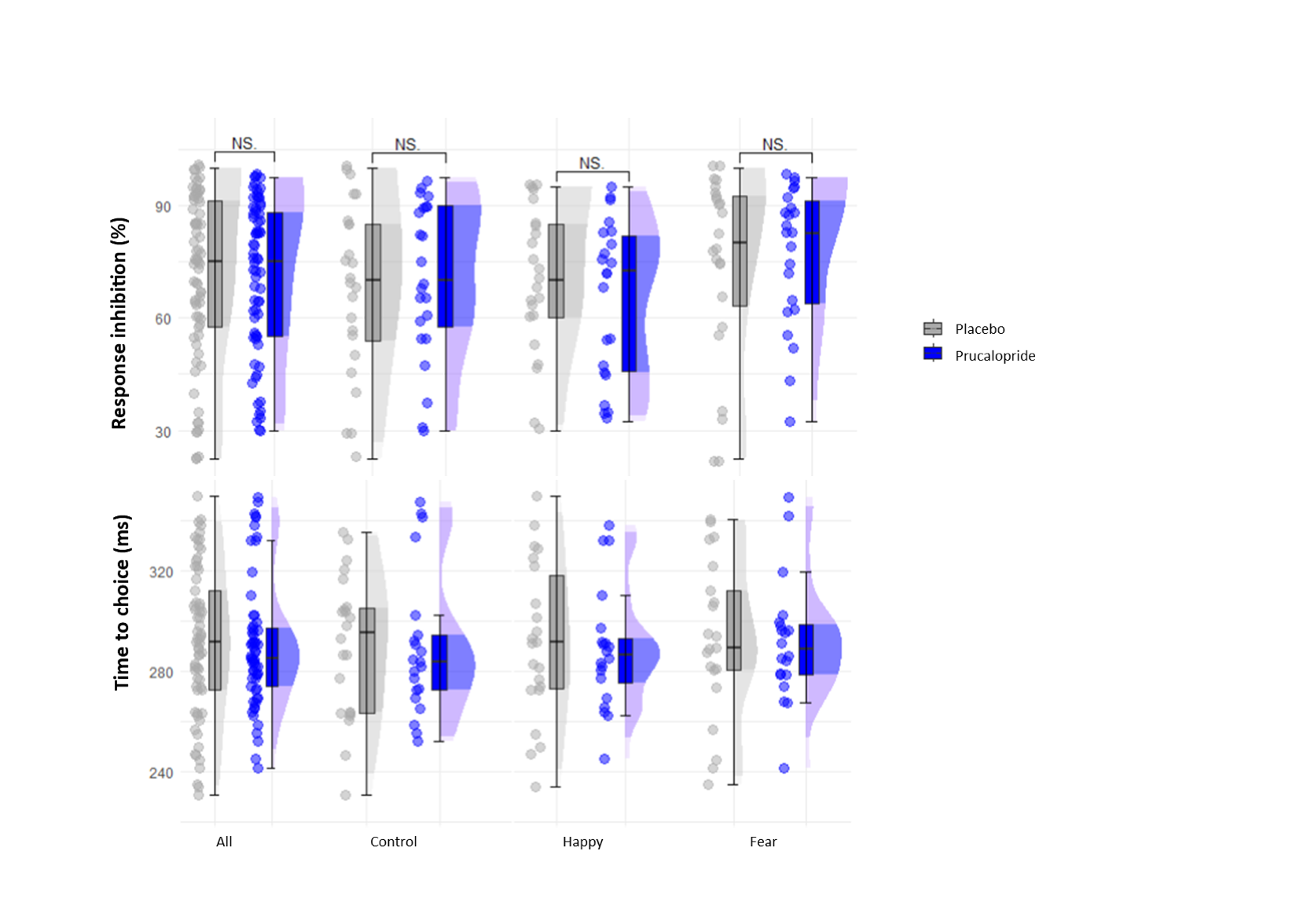

(A) Response inhibition (accuracy of no presses); (B) Time to choice (reaction time) by trial (all, control, happy, fear); Whiskers demonstrate 1.5 x interquartile range, central line represents the median, half violin plots represent the data distribution. NS = non-significant group differences

###### ***Figure S8D: Results of the EGNG task in prucalopride and placebo groups: log decision criterion (C)***

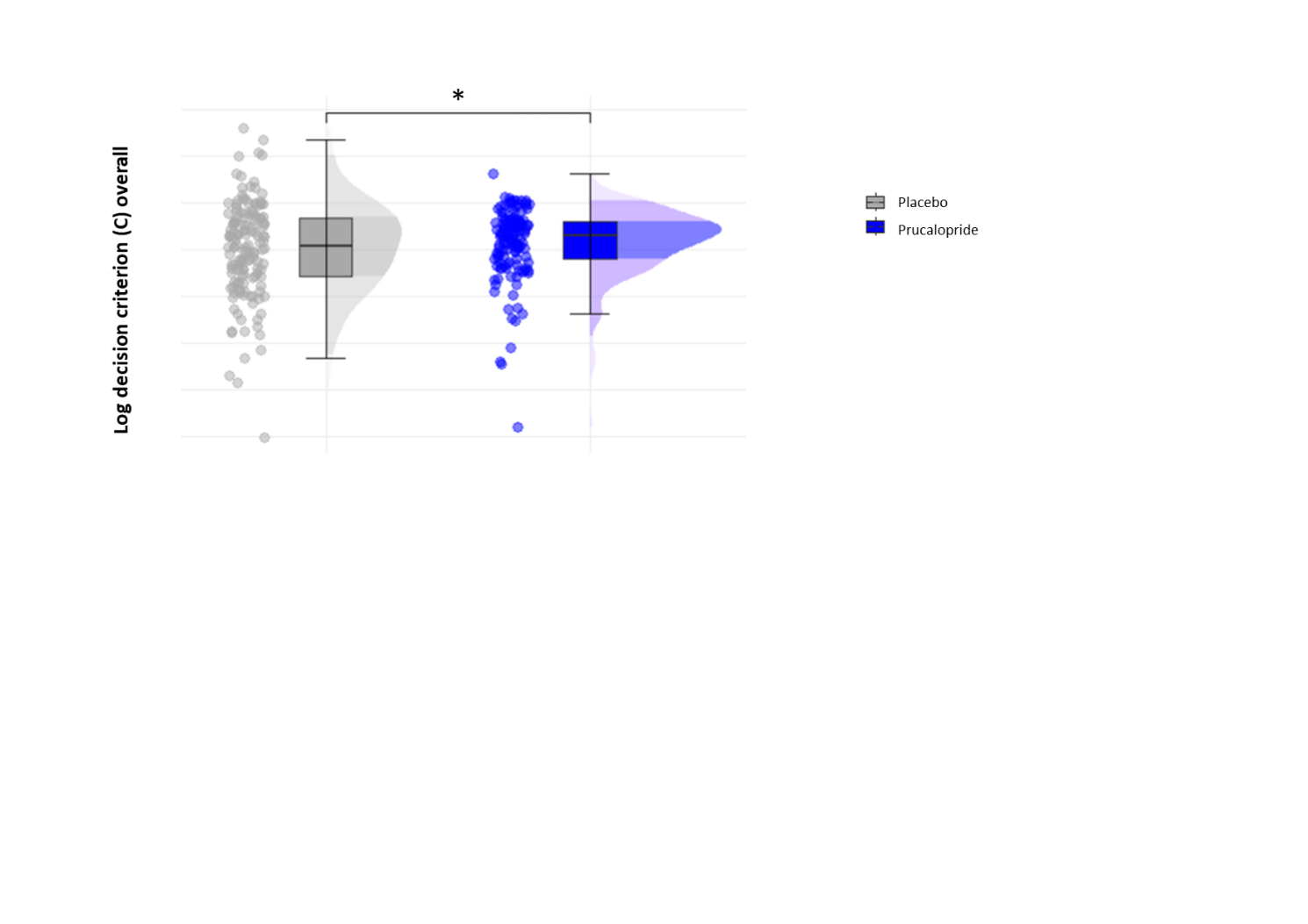

Whiskers demonstrate 1.5 x interquartile range, central line represents the median, half violin plots represent the data distribution. NS = non-significant group differences

###### Supplementary results for the EGNG

Choice impulsivity was not reduced when aversive emotional distractors were present (EMM = 3.52 ± 11.4, p = 0.76) compared with both control (EMM = 1.80 ± 11.4, p = 0.87) and positive emotional distractors (EMM = 1.54 ± 11.4, p = 0.89

*Sensitivity analyses for baseline mood and subjective cognition*

Accurate inhibition for no go presses was unchanged (ANCOVA main effect of group: F[1,42]=0.01, *p*=0.92)

Reaction time analysis for go trials was also unchanged (ANCOVA main effect of group: F[1,42] = 0.22, *p* = 0.89).

*Sensitivity analyses including two participants (1 placebo; 1 prucalopride) with reaction time trials outside of optimisation parameters (i.e.) trials with immediate responses although average responding within normal parameters (N = Placebo 25, Prucalopride 24)*

5-HT_4_R agonism was not associated with a change in response inhibition, measured by mean percentage of accurately withheld responses to no-go trials (ANCOVA main effect of group: F[1,46]=0.065, *p*=0.799, np2 < 0.01; Placebo mean (SD) 34.8 (8.4); Prucalopride mean (SD) 34.4 (7.9); all conditions EMM = 0.31 ± 1.84, *p* = 0.87, *d* = 0.02 [-0.21, 0.25] (Figure S5(A)). This was unchanged when we accounted for baseline scores of mood and subjective cognition (ANCOVA main effect of group: F[1,44]=0.065, *p*=0.801). There was also an absence of group by set shift interaction for accuracy of withheld presses (ANCOVA: F[1,340]=0.107, *p*=0.74). Further, there was no statistically significant group effect for go trial accuracy (ANCOVA main effect of group: F[1,46]=0.004, *p*=0.95) both when considering the control condition or affective interference (Figure S5(B)).

Prucalopride allocation did not affect choice impulsivity, indicated by no change in reaction time to choice for go trials, across all task conditions (ANCOVA main effect of group: F[1,46] = 0.20, p = 0.63; ηp2 < 0.01) (see Figure S5(C)). Specifically, choice impulsivity was not reduced when aversive emotional distractors were present (EMM = 3.52 ± 11.4, p = 0.76, d = 0.09 [-0.48, 0.65]) compared with both control (EMM = 1.80 ± 11.4, p = 0.87, d = 0.04 [-0.52, 0.61]) and positive emotional distractors (EMM = 1.54 ± 11.4, p = 0.89, d = 0.04 [-0.52, 0.60]). Reaction time analysis was also unchanged when we considered the role of mood and subjective cognition (ANCOVA main effect of group: F[1,44] = 0.22, *p* = 0.64).

Signal detection theory analyses was undertaken to determine if group differences in response inhibition were driven by perceptual decision-making. Prucalopride allocation did not affect caution regarding decision-making across task conditions (log criterion c; ANCOVA main effect of group: F[1,46] = 0.96, *p* = 0.33, η_p_^2^ < 0.01 [0.00, 0.00]; all conditions EMM = -0.015 ± 0.03, *p* = 0.58, *d* = -0.064 [-0.29, 0.16]) (see Figure S5(D)). There was also no statistically significant group effect on signal discriminability (d’prime: ANCOVA main effect of group: F[1,46] = 0.89, *p* = 0.35, η_p_^2^ = 0.02 [0.00, 0.20], all conditions EMM = 0.07 ± 0.12, *p* = 0.54, *d* = -0.064 [-0.29, 0.16]) or when considering the decision criterion (beta: ANCOVA main effect of group: F[1,46] = 0.81, *p* = 0.37, η_p_^2^ <0.01 [0.00, 0.13], all conditions EMM = -0.05 ± 0.09, *p* = 0.57, *d* = -0.066 [-0.30, 0.16].

*Drift diffusion modelling*

###### Table S10: Results from drift diffusion modelling

|  |  | **Boundary Separation** | **Non decision time** | **Initial Bias** | **Drift Bias** | **Drift Rate** |
| --- | --- | --- | --- | --- | --- | --- |
| **Mean (SD)** | ***Placebo*** | 1.13 (0.62) | 0.18 (0.07) | -1.13 (1.22) | 2.55 (3.85) | 3.68 (4.15) |
|  | ***Prucalopride*** | 0.94 (0.50) | 0.20 (0.06) | -0.94 (1.04) | 2.72 (2.62) | 2.88 (1.50) |
| **ANOVA** | ***Main effect (Group)*** | F(1,133)=3.05, p=0.08 | F(1,133)=2.79, p=0.10 | F(1,133)=1.70, p=0.19 | F(1,133)=0.01, p=0.92 | F(1,133)=2.08, p=0.15 |
|  | ***Group*trial type*** | F(1,133)=0.79, p=0.46 | F(1,133)=0.92, p=0.40 | F(1,133)=0.50, p=0.62 | F(1,133)=0.36, p=0.70 | F(1,133)=1.01, p=0.40 |

###### Figure S8E: Results from drift diffusion modelling

(A) **(B)**

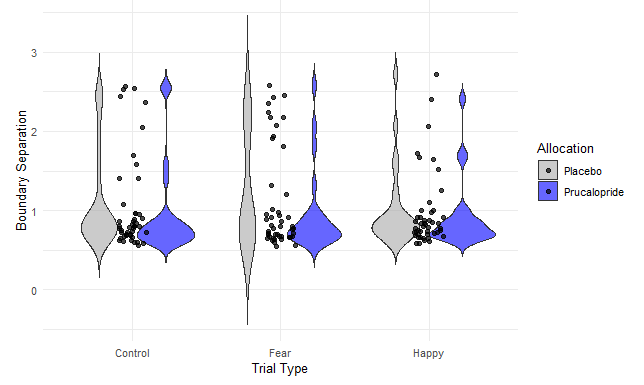

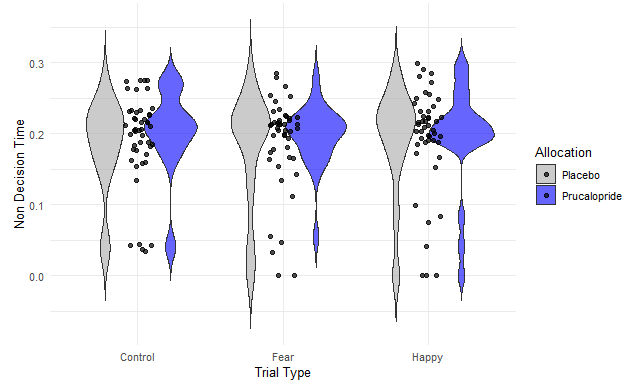

(C) **(D)**

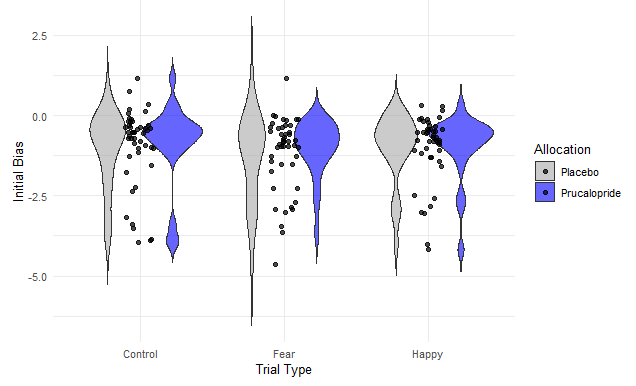

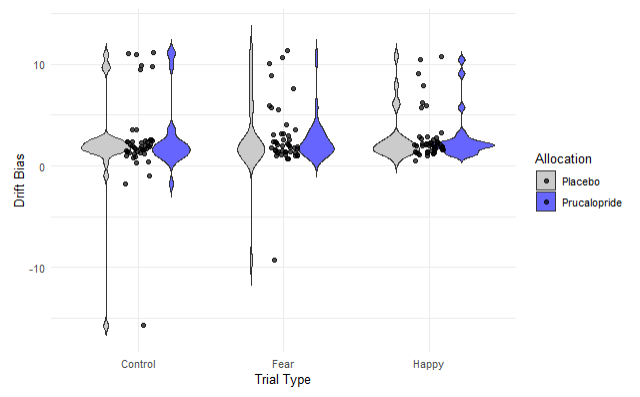

**(E)**

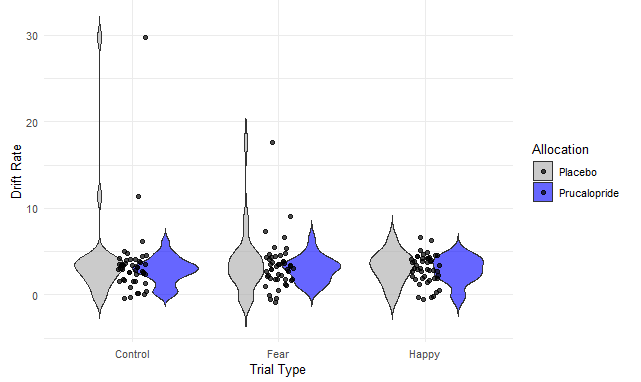

(A) Boundary Separation; (B) Non Decision Time; (C) Initial Bias; (D) Drift Bias; (E) Drift Rate. Violin plots showing spread of the data across groups.

#### ECAT

N = Placebo 25, Prucalopride 23

###### ***Table S11: Results for ECAT (accuracy and reaction time) by valence***

| **Measure** | **Condition** | **Prucalopride**  **(N=23, SD)** | **Placebo**  **(N=25, SD)** |
| --- | --- | --- | --- |
| ***Accuracy (%)*** | Positive | 94.1 (9.5) | 93.8 (11.4) |
|  | Negative | 93.9 (6.4) | 95.0 (5.4) |
| ***Reaction time (ms)*** | Positive | 750 (217) | 796 (169) |
|  | Negative | 798 (200) | 861 (158) |

###### Supplementary results for the ECAT

*Accuracy*

On the task evaluating speed to classify positive and negative personality characteristics (EREC), there was no significant main effect of group [F(1,46)=0.04, p=0.84, np2 < 0.01] or a significant interaction between the group and valence [F(1,46)=0.2, p=0.66, np2 < 0.01].

Results were very similar when PHQ9 scores and PDQ20 scores at baseline were incorporated into the model: main effect of group [F(1,44)=0.04, p=0.85, np2 < 0.01] or group × valence interaction [F(1,46)=0.2, p=0.65, np2 < 0.01], or when first language was included: main effect of group [F(1,45)=0.50, p=0.48, np2=0.01], accuracy × group interaction [F(1,45)=0.20, p=0.66, np2 < 0.01].

*Reaction time*

In terms of reaction time assessment, there was no significant main effect of group [F(1,46)=1.08, p=0.31, np2=0.02] or a significant interaction between the group and valence [F(1,46)=0.36, p=0.55, np2 < 0.01]; however there was a trend for the prucalopride group to be faster across both valences.

Results were very similar when PHQ9 scores and PDQ20 scores at baseline were incorporated into the model: main effect of group [F(1,44)=0.88, p=0.35, np2=0.02]] or group × valence interaction [F(1,44)=0.35, p=0.55, np2 < 0.01], or when first language was included: main effect of group [F(1,45)=0.49, p=0.49, np2=0.01], accuracy × group interaction [F(1,44)=0.39, p=0.53, np2 < 0.01].

###### ***Figure S9A and B: ECAT accuracy (hits) and reaction time by valence***

**(A)**

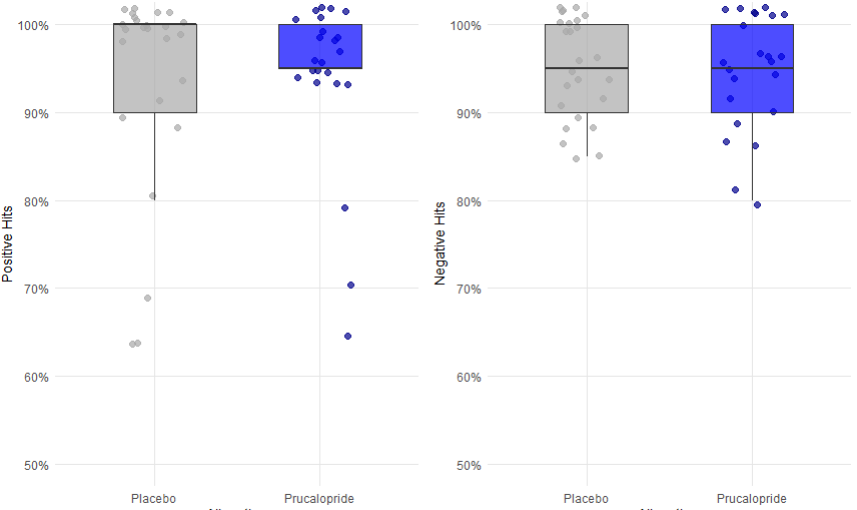

**(B)**

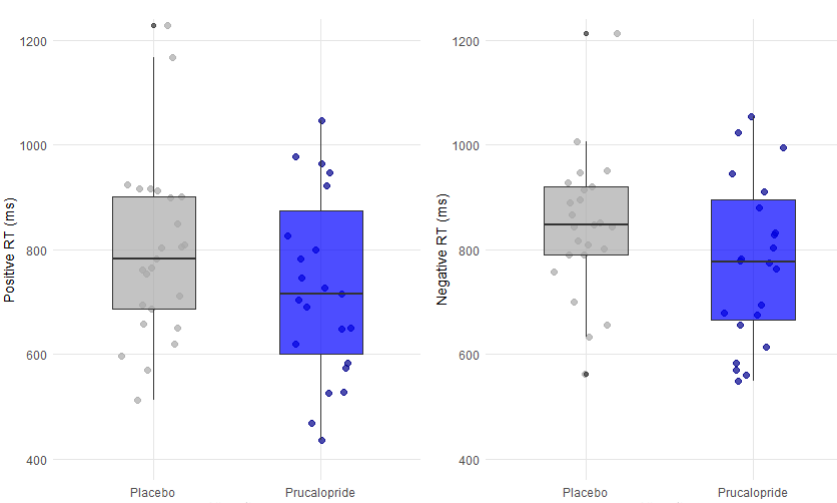

Whiskers demonstrate 1.5 x interquartile range, central line represents the median, data points represent the data distribution.

#### EREC

N = Placebo 25, Prucalopride 23

###### ***Table S12: Results for EREC (total items recalled and false alarms) by valence***

| **Measure** | **Condition** | **Prucalopride**  **(N=23, SD)** | **Placebo**  **(N=25, SD)** |
| --- | --- | --- | --- |
| ***Total items recalled*** | Positive | 2.47 (2.86) | 2.08 (2.52) |
|  | Negative | 1.57 (1.88) | 2.04 (2.65) |
| ***False alarms*** | Positive | 4.17 (3.68) | 4.12 (3.92) |
|  | Negative | 2.22 (2.80) | 2.52 (2.38) |

###### ***Figure S10: Box and whisker plots for EREC (total items recalled (hits) and false alarms) by valence***

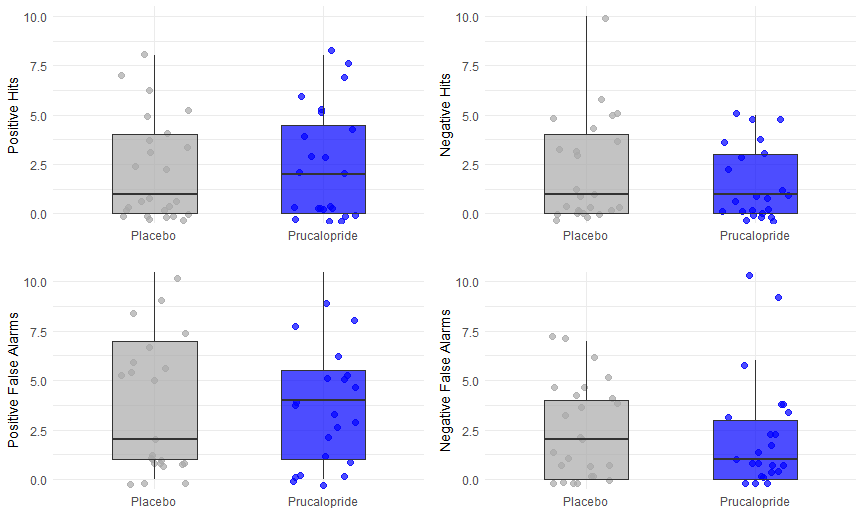

Whiskers demonstrate 1.5 x interquartile range, central line represents the media, data points represent the data distribution

###### Supplementary results for the EREC

On the emotional word recall task (EREC), there was no main effect of group (no significant interaction between the group and recall accuracy [F(1,46)=0.004, p=0.95, np2 < 0.01] or a three-way group × valence × accuracy interaction [F(1,46)=0.25, p = 0.61, np2 < 0.01]. This reflected no significant differences between groups on hits or false alarms for any valence (see Figure S6 and Table S4). Results were very similar when PHQ9 scores and PDQ20 scores at baseline were incorporated into the model: interaction between group and recall accuracy [F(1,40)=0.004, p=0.95, np2 < 0.01 ] or a three-way group × valence × accuracy interaction [F(1,40)=0.14, p=0.71, np2 < 0.01], and when we accounted for first language: main effect of group [F(1,43)=0.07, p=0.80, np2 < 0.01], interaction between group and recall accuracy [F(1,40) = 0.004, p = 0.95, np2 < 0.01], three-way group × valence × accuracy interaction [F(1,40)=0.22, p=0.64, np2 < 0.01].

#### EMEM

N = Placebo 25, Prucalopride 23

###### ***Table S13: Results for EMEM (accuracy and reaction time) by valence***

| **Measure** | **Condition** | **Prucalopride**  **(N=23)** | **Placebo**  **(N=25)** |
| --- | --- | --- | --- |
| ***Accuracy (%)*** | Positive | 16.0 (3.24) | 17.6 (2.33) |
|  | Negative | 12.3 (3.95) | 12.5 (5.87) |
| ***Reaction time (ms)*** | Positive | 975 (178) | 996 (224) |
|  | Negative | 1006 (184) | 1033 (214) |

###### ***Figure S11: Box and whisker plots for EMEM (total items recalled (hits) and false alarms) by valence***

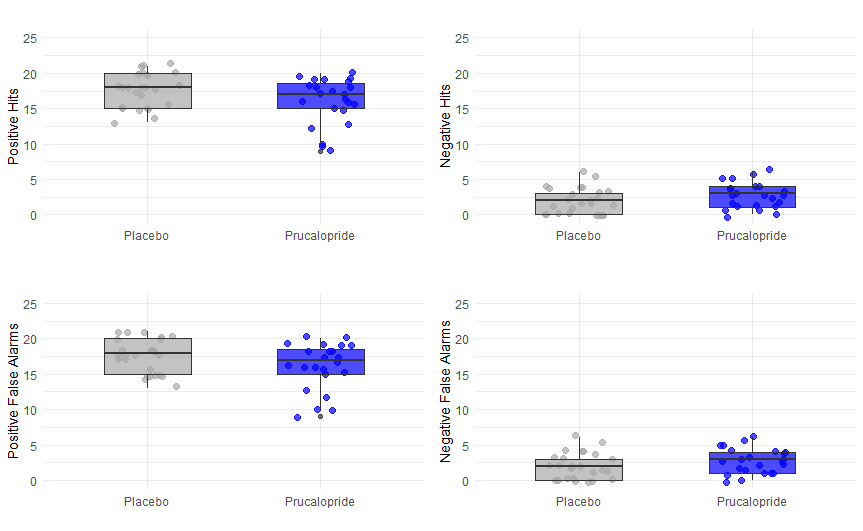

Whiskers demonstrate 1.5 x interquartile range, central line represents the media, data points represent the data distribution

###### Supplementary results for the EMEM

*Accuracy*

On the emotional word recognition task (EMEM) for performance, there was no main effect of group [F(1,46)=0.42, p=0.52, np2 < 0.01], and no significant accuracy × group interaction [F(1,46)=0.70, p=0.41, np2 < 0.01]. This was unchanged when PHQ9 scores and PDQ scores at baseline were included in the model: main effect of group [F(1,40)=0.48, p=0.49, np2 < 0.01], accuracy × group interaction [F(1,40)=0.69, p=0.41, np2 < 0.01], or when first language was included: main effect of group [F(1,43)=0.53, p=0.47, np2 = 0.01], accuracy × group interaction [F(1,43)=0.71, p=0.40, np2 < 0.01].

*Reaction time*

On the emotional word recognition task (EMEM) for reaction time, there was no main effect of group [F(1,46) = 0.18, p = 0.67, np2 < 0.01], and no significant accuracy × group interaction [F(1,46)=0.03, p=0.87, np2 < 0.01]. This was very similar when PHQ9 scores and PDQ scores at baseline were included in the model: main effect of group [F(1,44)=0.28, p=0.86, np2 < 0.01], accuracy × group interaction [F(1,44)=0.03, p=0.87, np2 < 0.01], or when first language was included: main effect of group [F(1,45)=0.20, p=0.66, np2 = 0.01], accuracy × group interaction [F(1,45)=0.04, p=0.87, np2 < 0.01].
